# Multinational Patterns of Second-line Anti-hyperglycemic Drug Initiation Across Cardiovascular Risk Groups: A Federated Pharmacoepidemiologic Evaluation in LEGEND-T2DM

**DOI:** 10.1101/2022.12.27.22283968

**Authors:** Rohan Khera, Lovedeep Singh Dhingra, Arya Aminorroaya, Kelly Li, Jin J Zhou, Faaizah Arshad, Clair Blacketer, Mary G Bowring, Fan Bu, Michael Cook, David A Dorr, Talita Duarte-Salles, Scott L DuVall, Thomas Falconer, Tina E French, Elizabeth E Hanchrow, Scott Horban, Wallis CY Lau, Jing Li, Yuntian Liu, Yuan Lu, Kenneth KC Man, Michael E Matheny, Nestoras Mathioudakis, Michael F McLemore, Evan Minty, Daniel R Morales, Paul Nagy, Akihiko Nishimura, Anna Ostropolets, Andrea Pistillo, Jose D Posada, Nicole Pratt, Carlen Reyes, Joseph Ross, Sarah L Seager, Nigam H Shah, Katherine R Simon, Eric YF Wan, Jianxiao Yang, Can Yin, Seng Chan You, Martijn J Schuemie, Patrick B Ryan, George Hripcsak, Harlan M Krumholz, Marc A Suchard

**Author notes:** **Correspondence to:** Marc A Suchard, MD, PhD, Departments of Biomathematics, Biostatistics and Human Genetics, 6558 Gonda Building, 695 Charles E. Young Drive, South Los Angeles, CA 90095-1766; @suchard_group.

## Abstract

**Objectives:** To assess the uptake of second-line antihyperglycemic agents among patients with type-2 diabetes mellitus (T2DM) receiving metformin.

**Design:** Serial cross-sectional study (2011-2021).

**Setting:** Ten US and seven non-US electronic health record and administrative claims databases in the Observational Health Data Sciences and Informatics network.

**Participants:** 4.8 million patients with T2DM receiving metformin.

**Main Outcomes Measures:** Calendar-year trends in the proportional initiation of second-line antihyperglycemic agents, glucagon-like peptide-1 receptor agonists (GLP-1 RAs), sodium-glucose cotransporter 2 inhibitors (SGLT2is), dipeptidyl peptidase-4 inhibitors, and sulfonylureas, for each database. We also evaluated the relative drug class-level uptake across cardiovascular risk groups.

**Results:** We identified 4.6 million patients with T2DM in US databases, 61,382 from Spain, 32,442 from Germany, 25,173 from the UK, 13,270 from France, 5,580 from Scotland, 4,614 from Hong Kong, and 2,322 from Australia. During 2011-2021, the combined proportional initiation of cardioprotective antihyperglycemic agents, GLP-1 RAs and SGLT2is, increased across all data sources, with the combined initiation of these drugs as second-line agents in 2021 ranging from 35.2% to 68.2% in the US databases, 15.4% in France, 34.7% in Spain, 50.1% in Germany, and 54.8% in Scotland. From 2016 to 2021, in some US and non-US databases, uptake of GLP-1 RAs and SGLT2is increased more significantly among populations without cardiovascular disease compared to those with established cardiovascular disease, without any data source providing evidence of a greater increase in their uptake in the populations with cardiovascular disease.

**Conclusions:** Despite the increase in overall uptake of cardioprotective antihyperglycemic agents as second-line treatment for T2DM, their uptake was lower in patients with cardiovascular disease over the last decade. A strategy to ensure medication use concordant with guideline recommendations is essential to improve outcomes of patients with T2DM.

## BACKGROUND

The management of type 2 diabetes mellitus (T2DM) has evolved over the last decade with both the introduction of novel drug agents, as well as an emphasis on lowering cardiovascular and renal risks. In large randomized clinical trials with T2DM patients, glucagon-like peptide-1 receptor agonists (GLP-1 RAs) and sodium-glucose cotransporter 2 inhibitors (SGLT2is) have strong evidence not only of addressing hyperglycemia but also of improving cardiovascular risk in high risk populations,[1–5] with SGLT2is additionally reducing the progression of renal disease.[1–3] Consequently, international clinical practice guidelines increasingly recognize the evolution of second-line agents as a treatment option for diabetes,[6] favoring the use of GLP-1 RAs in over a third and SGLT2is in over half of all patients with T2DM.[7]

Despite clinical trial and real-world data evidence supporting the benefits of GLP-1 RAs, since 2017, and of SGLT2is, since 2016, the actual uptake of these drugs continues to lag.[7–12] Further, studies characterizing patterns of use have exclusively focused on prevalent use, and US-based studies have focused on single-payers or small populations included in national surveys. These assessments likely do not accurately capture the uptake patterns for novel therapies, for which both the uptake and the use are likely to grow over time. Moreover, the cost of these drugs and their coverage through health insurance programs varies across healthcare systems and countries.[13–16]

An appraisal of the uptake of GLP-1 RAs and SGLT2is as second-line therapy among those escalated from metformin monotherapy is critical. This is particularly relevant as an assessment of their initiation relative to other second-line agents, namely, dipeptidyl peptidase-4 inhibitors (DPP-4is) and sulfonylureas (SUs) that have been available for longer, but lack cardioprotective or renoprotective effects.[17–20] In a large, multinational study, we describe patterns of initiation of four key second-line agents— GLP-1 RAs, SGLT2is, DPP-4is, and SUs — during escalation from metformin monotherapy, overall and across clinical and demographic subgroups.

## METHODS

### Study Overview

This study represents a federated pharmacoepidemiology analysis among T2DM patient records from a multinational consortium of data sources all mapped to the Observational Medical Outcomes Partnership (OMOP) Common Data Model (CDM).[21] We defined a cohort of T2DM patients receiving metformin therapy who were initiated on second-line anti-hyperglycemic agents and evaluated patterns of uptake of traditionally second-line anti-hyperglycemic agents with and without known cardioprotective effects, across patients spanning the cardiovascular risk spectrum.

### Data Sources

We identified participating data sources in the Large-scale Evidence Generation and Evaluation across a Network of Databases for Type 2 Diabetes Mellitus (LEGEND-T2DM) initiative. LEGEND-T2DM has been previously described.[22] Briefly, LEGEND-T2DM is a series of systematic, large-scale observational studies of real-world characterization of second-line anti-hyperglycemic agents. Of these, this present study is based on 17 real-world data sources, spanning administrative claims and electronic health record (EHR) databases, including six national-level and four health-system datasets from the US, and data sources from Spain, Germany, UK, France, Scotland, Hong Kong, and Australia. Further details about the data sources are included in **Table 1** and **Supplemental Table S1**. Patient records spanned the last decade (2011 - 2021) during which several second-line anti-hyperglycemic agents have been introduced. The most recent data available across data sources varied from 2019 through 2021 (**Table 1**). All patient records were standardized to the Observational Health Data Sciences and Informatics (OHDSI)’s OMOP CDM version 5, mapping international coding systems into standard vocabulary concepts.[23] These data sources have previously been leveraged in OHDSI studies.[24–26]

**Table 1.** Description of Databases from the Observational Health Data Sciences and Informatics Network Included in the Study.

| Name of Database | Abbreviation | Type of Data | Country of Origin | Years of Exposure Included | Number of Participants |
| --- | --- | --- | --- | --- | --- |
| **US National Databases** |  |  |  |  |  |
| IBM MarketScan® Commercial Claims and Encounters Data (CCAE) | CCAE | Claims Data | USA | 2011-2021 | 265,874 |
| IBM Health MarketScan® Multi-State Medicaid Database | MDCD | Claims Data | USA | 2011-2020 | 40,064 |
| IBM Health MarketScan® Medicare Supplemental and Coordination of Benefits Database | MDCR | Claims Data | USA | 2011-2021 | 43,857 |
| Optum Clinformatics Extended Data Mart - Date of Death (DOD) | OCEDM | Claims Data | USA | 2011-2021 | 211,877 |
| Optum® de-identified Electronic Health Record Dataset | OEHR | Claims Data | USA | 2011-2021 | 299,008 |
| US Open Claims | USOC | Claims Data | USA | 2000-2021 | 3,521,191 |
| **US Health System Databases** |  |  |  |  |  |
| Columbia University Irving Medical Center | CUIMC | EHR Data | USA | 2011-2021 | 4,561 |
| Johns Hopkins Medicine | JHM | EHR Data | USA | 2016-2021 | 3,759 |
| Stanford Medicine | STARR | EHR Data | USA | 2011-2021 | 2,993 |
| Department of Veterans Affairs Healthcare System | VA | EHR Data | USA | 2011-2021 | 230,019 |
| **Non-US Databases** |  |  |  |  |  |
| Australia Longitudinal Patient Database Practice Profile | ALPD | EHR Data | Australia | 2012-2021 | 2,322 |
| France Longitudinal Patient Database | FLPD | EHR Data | France | 2012-2021 | 13,270 |
| Germany Disease Analyser | GDA | EHR Data | Germany | 1992-2021 | 32,442 |
| Health Informatics Centre at the University of Dundee | HIC | EHR Data | Scotland | 2011-2021 | 5,580 |
| HKHA - Hong Kong Hospital Authority | HKHA | EHR Data | Hong Kong | 2011-2018 | 4,614 |
| UK-IQVIA Medical Research Data | IMRD | EHR Data | United Kingdom | 2011-2019 | 25,173 |
| Information System for Research in Primary Care | SIDIAP | EHR Data | Spain | 2011-2021 | 61,382 |

The US populations included those commercially and publicly insured, enriched for older individuals (Medicare [MDCR], Veterans Health Administration [VA]), lower socioeconomic status (Managed Medicaid [MDCD]), and racially diverse populations (>20% Black or African American in the VA, and 8% in Columbia University Irving Medical Center [CUIMC]). The study was designed at a data source level and followed federated analytic principles, so the same patients may be represented in more than one data source, particularly in the US. All data sources received institutional review board approval or exemption for their participation in LEGEND-T2DM. The study is reported according to the Strengthening the Reporting of Observational Studies in Epidemiology (STROBE) reporting guidelines.[27]

### Study Population

We included all adult subjects (age ≥18 years) traditionally included in second-line T2DM agent exposure cohorts, as described in the LEGEND-T2DM study protocol.[22] Broadly, these cohorts consisted of T2DM patients who had prior metformin monotherapy and initiated second-line treatment with one of the 22 drug ingredients that comprise the GLP-1 RAs, SGLT2is, DPP-4is, and SUs drug classes (**Supplemental Table S2**). We did not consider thiazolidinediones given their known association with a risk of heart failure and bladder cancer.[28,29] The study population included patients with and without established cardiovascular disease (CVD) based on the previously developed and validated definition for risk stratification among new users of second-line T2DM agents.[30] The methodology of cohort definition is detailed in the **Supplemental Methods**.

### Study Exposures and Outcomes

This study evaluated changes in patterns of second-line anti-hyperglycemic initiation over time. We used calendar years as the exposure, with the years in the study that were specific to each cohort (outlined in **Table 1**). The outcome was the incidence of second-line antihyperglycemic agents use among all individuals who were already receiving treatment with metformin.

### Study Covariates

Study covariates were drawn from the broad set of characteristics outlined in the cohort characterization tool stack in OHDSI.[31] We defined cohort demographics including age, sex, and race. The clinical characteristics were defined by standard OMOP concepts for diseases and procedures, spanning all body systems, representing 33 covariates. A team of clinicians verified the covariates included for presentation in the study to focus on those relevant to the management of diabetes, spanning domains of cardiovascular risk factors, established CVD, and kidney disease.

### Statistical Analysis

We evaluated the trend of yearly incident use of all four second-line antihyperglycemic agent classes across 17 databases. For each year, we excluded a database for analyses if the number of subjects in the database was less than 100. The number of subjects in the databases for each year is provided in **Supplemental Table S3**. Given both protective effects of GLP-1 RAs and SGLT2is on cardiovascular outcomes, we further performed a stratified analysis among individuals with and without established CVD (**Supplemental Methods**). In order to calculate the annual changes of the incidence rates for second-line antihyperglycemic drugs initiation from 2016 to 2021, we fitted linear regression models to the data using incidence rate as the dependent variable and year, coded as 1 to 6, as the independent variable. The annual change was reported as the point estimate of the slope (95% confidence interval). We compared the annual changes between patients with and without CVD for each second-line agent using the interaction term of CVD status and year in analysis of covariance (ANCOVA) models. We developed an interactive webpage to allow exploration of the cohorts included in LEGEND-T2DM.[32]

### Patient and Public Involvement

Patients and/or public were not specifically involved in the development of research hypothesis or the outcome measures, or in the design and implementation of the study due to the federated approach of the study.

## RESULTS

### Cohort Characteristics

LEGEND-T2DM included a total of over 4.8 million patients with T2DM across all cohorts, representing individuals initiating one of the four second-line anti-hyperglycemic agents between 2011-2021 (**Table 1**). This included 4.6 million T2DM patients initiating second-line therapy across US-based databases and 145,000 from non-US databases. Among the US databases, the US Open Claims contributed the maximum of 3.5 million patient records. The non-US data includes 61,382 patient records from Spain, 32,442 from Germany, 25,173 from the UK, 13,270 from France, 5,580 from Scotland, 4,614 from Hong Kong, and 2,322 from Australia.

### Patient Characteristics

Patients with T2DM who initiated GLP-1 RA second-line were more frequently female, while those initiating SGLT2is were more frequently male. Patients who were prescribed GLP-1 RA as the second-line therapy for T2DM had a lower prevalence of CVD, including ischemic heart disease (IHD), cerebrovascular disease, and heart failure. For instance, 2.7% of GLP-1 RAs users in US Open Claims database had IHD, compared with 4.1% of SGLT2i users, 4.1% of DPP-4i users, and 4.1% SU of users (**Supplemental Tables S4-S7**).

Similarly, for the IBM Health MarketScan® Commercial Claims and Encounters Database (CCAE), 3.6% of the GLP-1 RA users had IHD, compared to 4.3% of SGLT2i users, 3.9% of DDP-4i users, and 4.3% of SU users. Both in the US and non-US databases, fewer patients initiating GLP-1 RAs and SGLT2is had renal impairment at baseline. For instance, in US Open Claims, 4.1% of GLP-1 RA and SGLT2i users had renal impairment, compared with 6.5% of DPP-4i users, and 6.7% of SU users. In the Information System for Research in Primary Care (SIDIAP) dataset from Spain, 1.5% each of patients prescribed GLP-1 RAs and SGLT2is had renal impairment, compared with 3.9% of DPP-4i users, and 1.7% of SU users (**Supplemental Tables S8-S11**).

### Incident Use of Second-Line Antihyperglycemic Drugs in 2021 Across Cohorts

In 2021, the choice of the prescribed second-line antihyperglycemic agent varied among different US databases. The combined incident use of cardioprotective agents, GLP-1 RAs and SGLT2is, ranged from 35.2% in Veterans Affairs Health System (VA) to 68.2% in Columbia University Irving Medical Center (CUIMC). The incident use of DDP-4is ranged from 14.5% in Stanford (STARR) to 23.5% in the VA. In contrast, SU incident use ranged from 11.1% in CUIMC to 41.3% in the VA (**Figures 1 and 2**).

**Figure 1.**
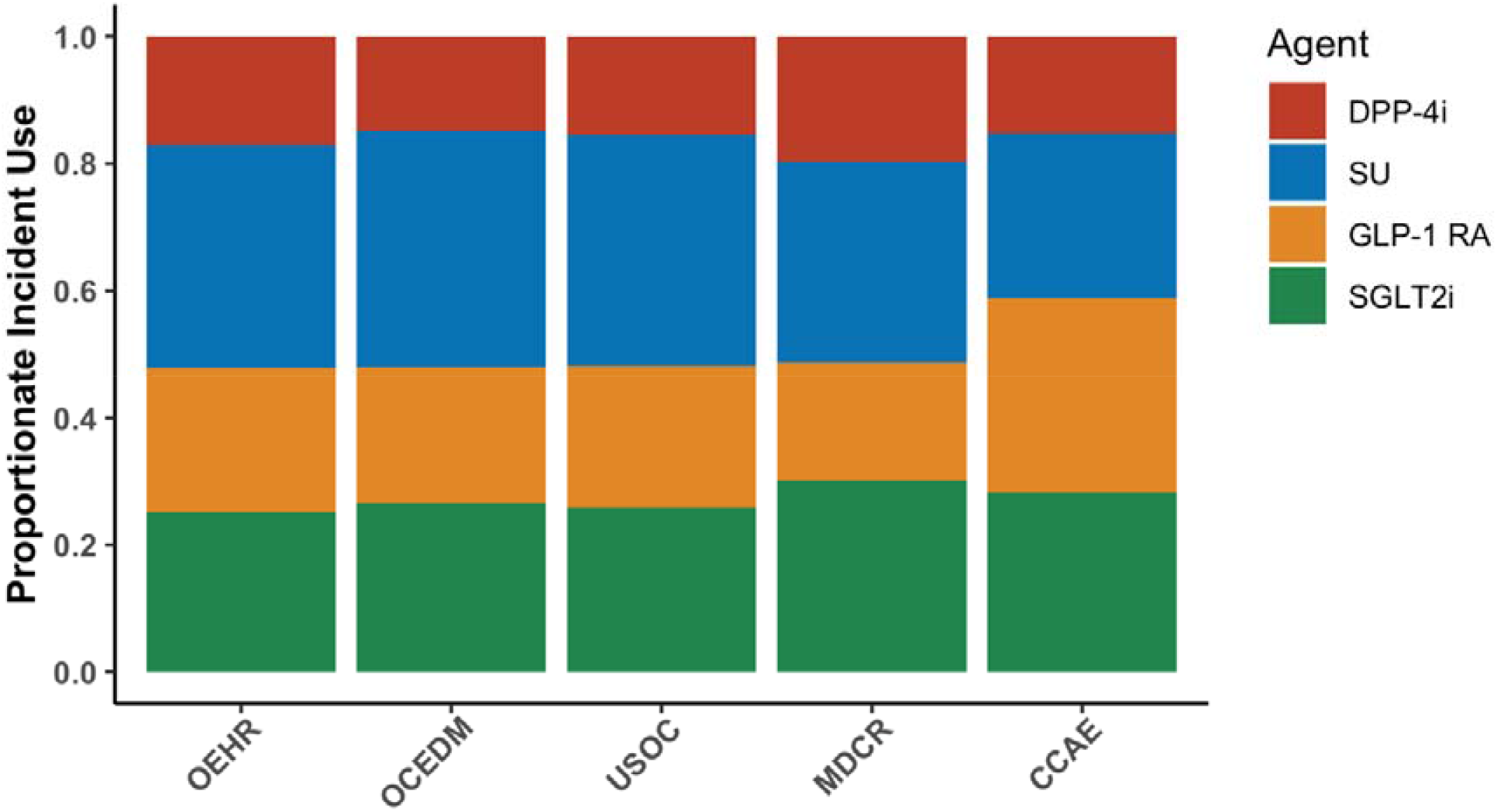
Proportional Incident Use of Second-Line Antihyperglycemic Agents in United States National Databases in 2021. **Abbreviations:** CCAE - IBM MarketScan® Commercial Claims and Encounters Data (CCAE), DPP-4i - Dipeptidyl Peptidase-4 Inhibitors, GLP-1 RA - Glucagon-like Peptide-1 Receptor Agonist, MDCR - IBM Health MarketScan® Medicare Supplemental and Coordination of Benefits Database, OCEDM - Optum Clinformatics Extended Data Mart - Date of Death (DOD), OEHR - Optum© de-identified Electronic Health Record Dataset, SGLT2i - Sodium-Glucose Cotransporter 2 Inhibitor, SU - Sulfonylurea, USOC - United States Open Claims

**Figure 2.**
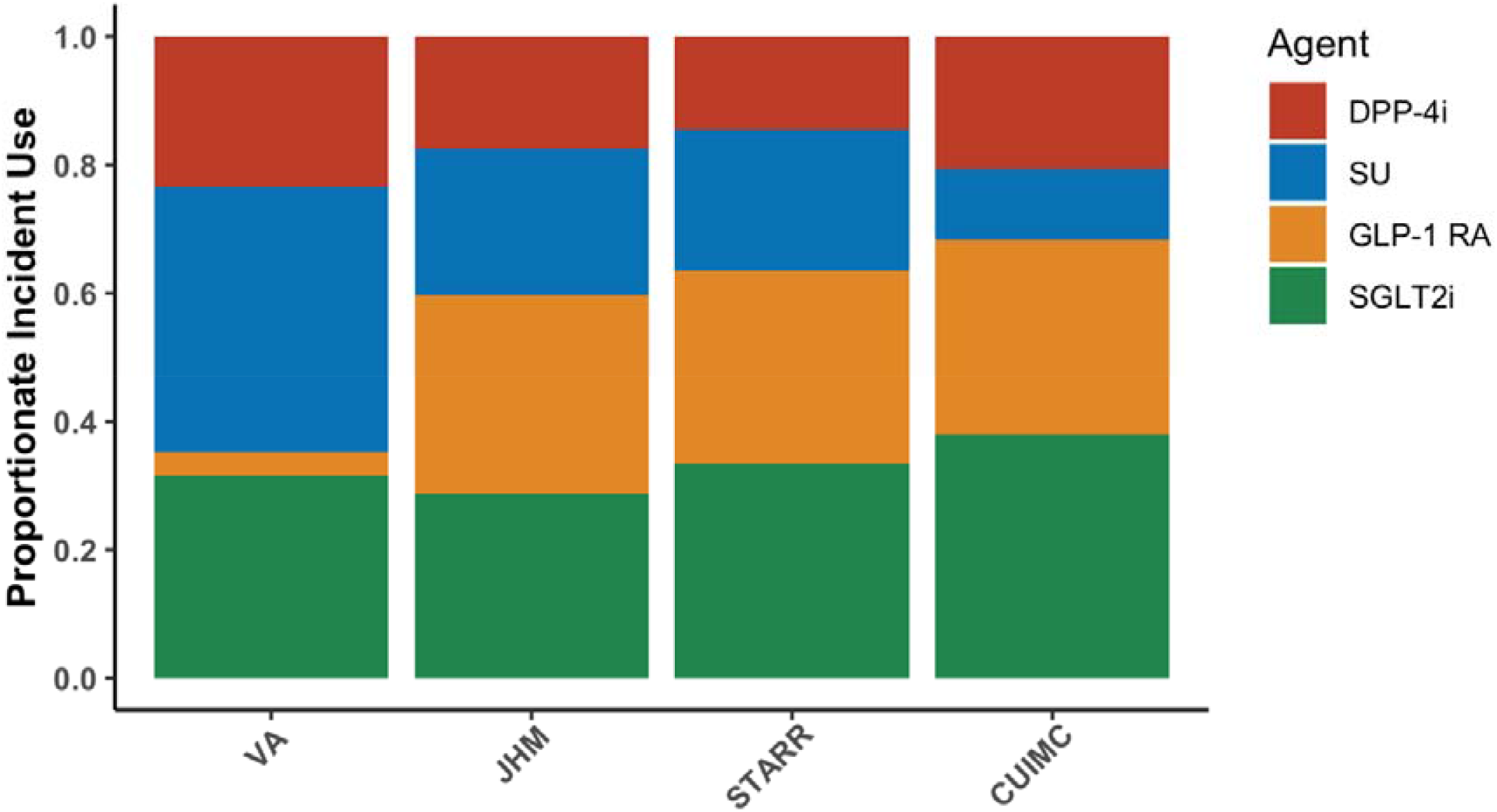
Proportional Incident Use of Second-Line Antihyperglycemic Agents in United States Health System Databases in 2021. **Abbreviations:** CUIMC - Columbia University Irving Medical Center, DPP-4i - Dipeptidyl Peptidase-4 Inhibitors, GLP-1 RA - Glucagon-like Peptide-1 Receptor Agonist, JHM - Johns Hopkins Medicine, SGLT2i - Sodium-Glucose Cotransporter 2 Inhibitor, STARR - Stanford Medicine, SU - Sulfonylurea, VA - Department of Veterans Affairs Healthcare System

Among the non-US databases, in 2021, the combined incident use of cardioprotective agents differed widely, ranging from 15.4% in France up to 54.8% in Scotland (**Figure 3**). Incident use of DPP-4is was greater in other countries than in the US, ranging from 44.2% in Scotland to 77.0% in France. In contrast, the incident use of SUs was lesser across the non-US databases as compared to the US databases, ranging from 1% in Scotland to 7.5% in France. The incident use of various antihyperglycemic agents in 2020 is shown in **Supplemental Figures S1-S3**.

**Figure 3.**
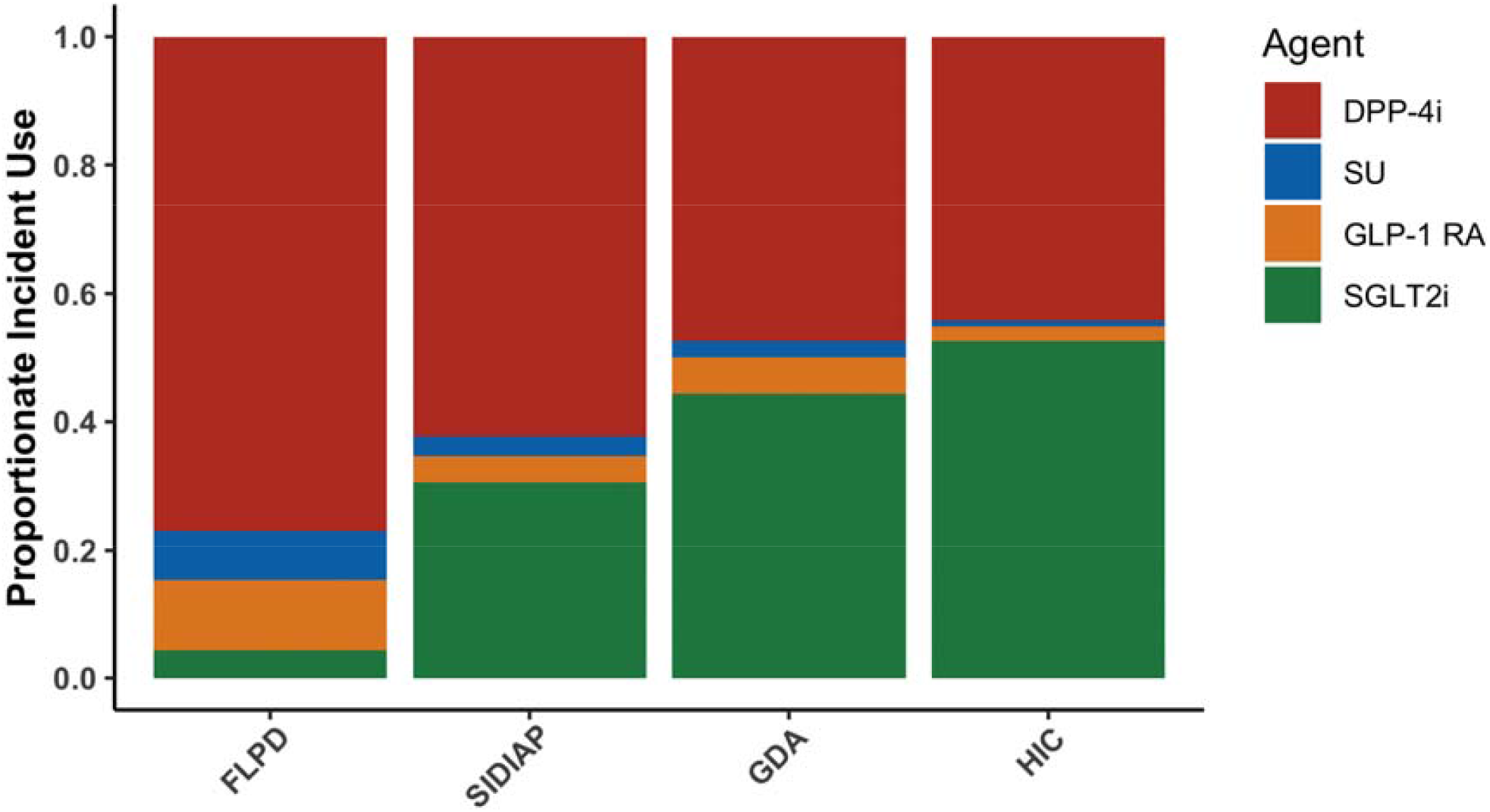
Proportional Incident Use of Second-Line Antihyperglycemic Agents in non-United States Databases in 2021. **Abbreviations:** ALPD - Australia Longitudinal Patient Database Practice Profile, DPP-4i - Dipeptidyl Peptidase-4 Inhibitors, FLPD - France Longitudinal Patient Database, GDA - Germany Disease Analyser, GLP-1 RA - Glucagon-like Peptide-1 Receptor Agonist, HIC - Health Informatics Centre at the University of Dundee, HKHA - Hong Kong Hospital Authority, SGLT2i - Sodium-Glucose Cotransporter 2 Inhibitor, SIDIAP - Information System for Research in Primary Care, SU – Sulfonylurea

### Trends in Uptake of Second-Line Antihyperglycemic Drug Use Across Study Cohorts

The proportion of second-line antihyperglycemic drug uptake varied across cohorts. Between 2011 and 2021, the initiation of GLP-1 RAs as the second-line antihyperglycemic agent increased across all US national data sources, from no measured initiation in 2011 to 18.5% in 2021 in the IBM Health MarketScan® Medicare (MDCR) population, and 30.5% in CCAE (**Supplemental Figure S4**).

Similarly, the uptake of SGLT2is in the US national databases increased from no uptake in 2011 across data sources to 25.2% in 2021 in the Optum© de-identified Electronic Health Record Dataset (OEHR) and 30.2% in the Medicare population. The VA had the lowest proportionate incident use of the cardioprotective antihyperglycemic agents in the US, driven predominantly by the low use of GLP-1 RAs (**Supplemental Figures S5**). The uptake of SGLT2is in the non-US databases increased from no uptake in 2011 to 4.4% in France and up to 52.6% in Scotland by 2021. Throughout the study period, there was limited use of GLP-1 RAs in Australia. However, among the non-US databases available, the use of GLP-1 RAs increased most in France, increasing to 11.1% in 2021 (**Supplemental Figure S6**).

From 2016 to 2021, the annualized increase in the combined incident use of GLP-1 RAs and SGLT2is was 10.6% per year in CUIMC, and 6.2% per year in US Open Claims database. In Spain, the annualized increase from 2016 to 2021 was 4.3% per year, compared to 2.7% per year in France, and 5.2% per year in Scotland.

### Second-Line Antihyperglycemic Drug Use Across Cardiovascular Risk Groups

The uptake of GLP-1 RAs in patients with established CVD in US national databases increased consistently from no incident use across databases in 2011 to 15.7% in Medicare (MDCR) patients and up to 28.0% in the CCAE population in 2021 (**Figure 4**). In contrast, the incident use of GLP-1 RA in patients without established CVD increased from no uptake in 2011 to 22.3% in MDCR patients and up to 38.0% in CUIMC patients in 2021 (**Figure 5**). Meanwhile, the incident use of SGLT2is in the patients with established CVD, in the same period, reached 28.7% in Optum Clinformatics Extended DataMart (OCEDM) and 46.0% in CUIMC (**Figure 6**). In patients without CVD, the increase in SGLT2is uptake was up to 23.3% in Optum© de-identified Electronic Health Record Dataset (OEHR) and up to 32.7% at Stanford Medicine (STARR) (**Figure 7**).

**Figure 4.**
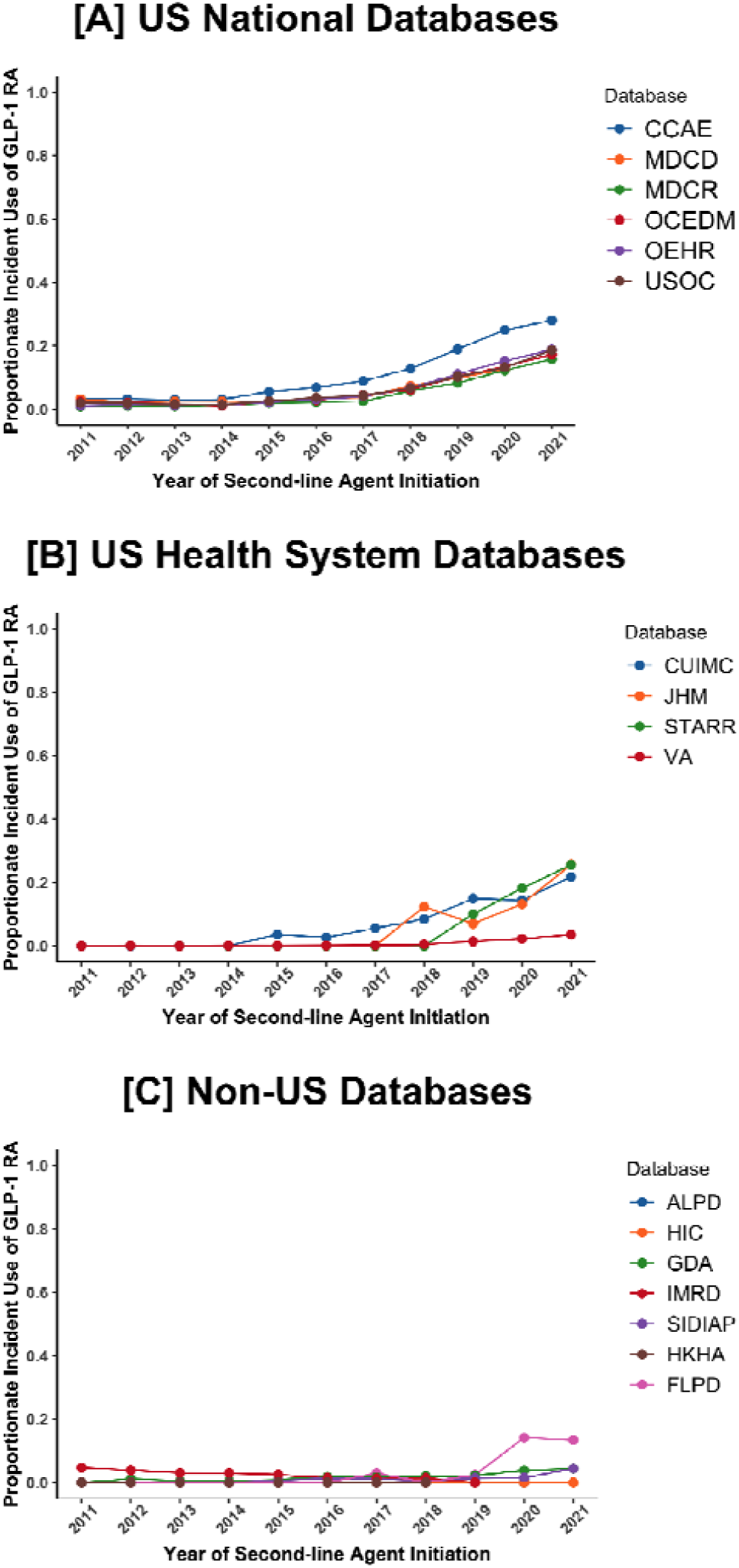
Proportional First Incident Use of Glucagon-like Peptide-1 Receptor Agonists as Second-line Therapy after Metformin in Patients with Established Cardiovascular Disease. **Abbreviations:** ALPD - Australia Longitudinal Patient Database Practice Profile, CCAE - IBM MarketScan® Commercial Claims and Encounters Data (CCAE), CUIMC - Columbia University Irving Medical Center, FLPD - France Longitudinal Patient Database, GDA - Germany Disease Analyser, GLP-1 RA - Glucagon-like Peptide-1 Receptor Agonist, HIC - Health Informatics Centre at the University of Dundee, HKHA - Hong Kong Hospital Authority, IMRD - UK-IQVIA Medical Research Data, JHM - Johns Hopkins Medicine, MDCD - IBM Health MarketScan® Multi-State Medicaid Database, MDCR - IBM Health MarketScan® Medicare Supplemental and Coordination of Benefits Database, OCEDM - Optum Clinformatics Extended Data Mart - Date of Death (DOD), OEHR - Optum© de-identified Electronic Health Record Dataset, SIDIAP - Information System for Research in Primary Care, STARR - Stanford Medicine, USOC - United States Open Claims, VA - Department of Veterans Affairs Healthcare System

**Figure 5.**
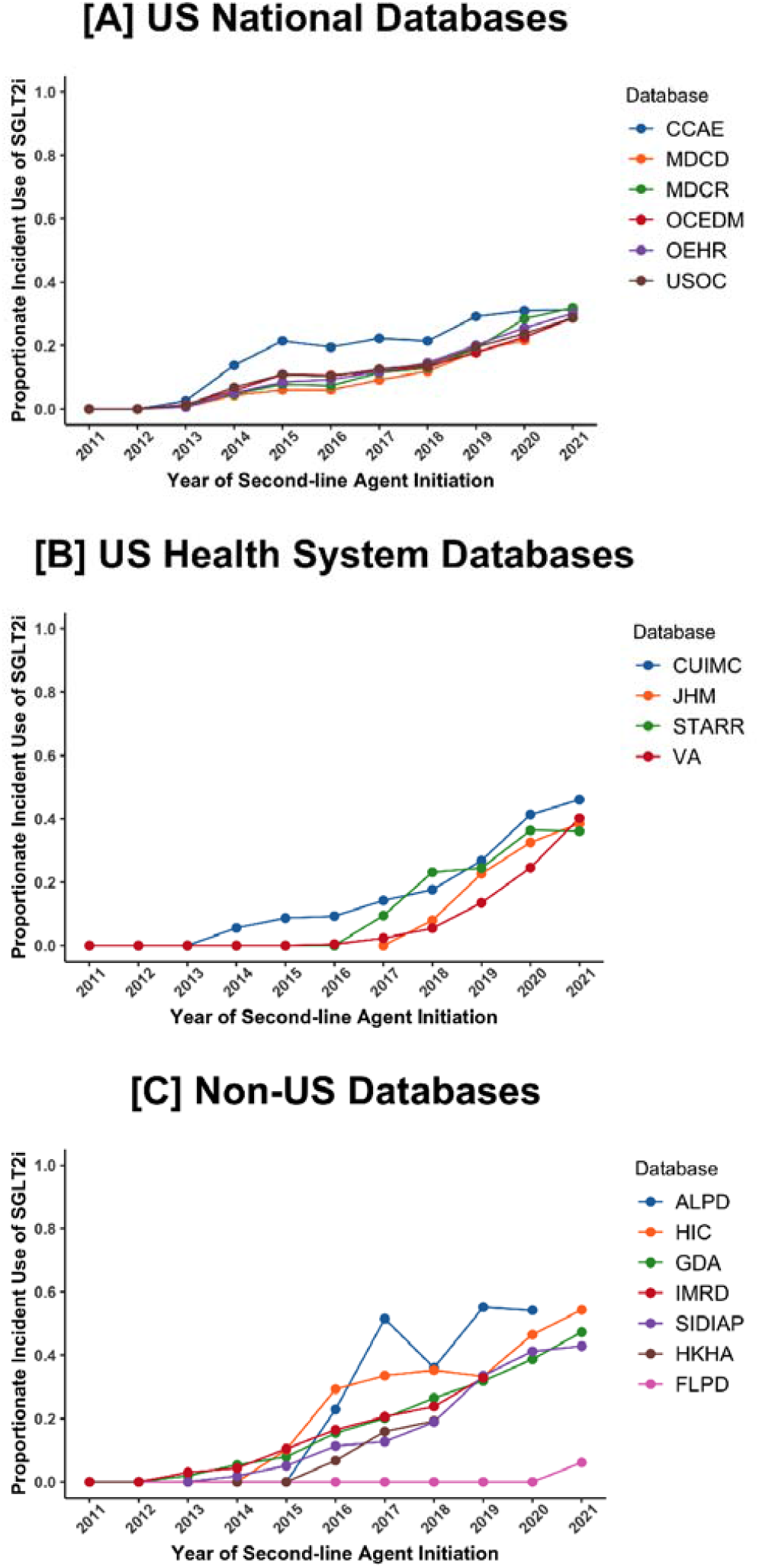
Proportional First Incident Use of Glucagon-like Peptide-1 Receptor Agonists as Second-line Therapy after Metformin in Patients without Established Cardiovascular Disease. **Abbreviations:** ALPD - Australia Longitudinal Patient Database Practice Profile, CCAE - IBM MarketScan® Commercial Claims and Encounters Data (CCAE), CUIMC - Columbia University Irving Medical Center, FLPD - France Longitudinal Patient Database, GDA - Germany Disease Analyser, GLP-1 RA - Glucagon-like Peptide-1 Receptor Agonist, HIC – Health Informatics Centre at the University of Dundee, HKHA - Hong Kong Hospital Authority, IMRD – UK-IQVIA Medical Research Data, JHM – Johns Hopkins Medicine, MDCD – IBM Health MarketScan® Multi-State Medicaid Database, MDCR – IBM Health MarketScan® Medicare Supplemental and Coordination of Benefits Database, OCEDM – Optum Clinformatics Extended Data Mart – Date of Death (DOD), OEHR – Optum© de-identified Electronic Health Record Dataset, SIDIAP – Information System for Research in Primary Care, STARR – Stanford Medicine, USOC – United States Open Claims, VA – Department of Veterans Affairs Healthcare System

**Figure 6.**
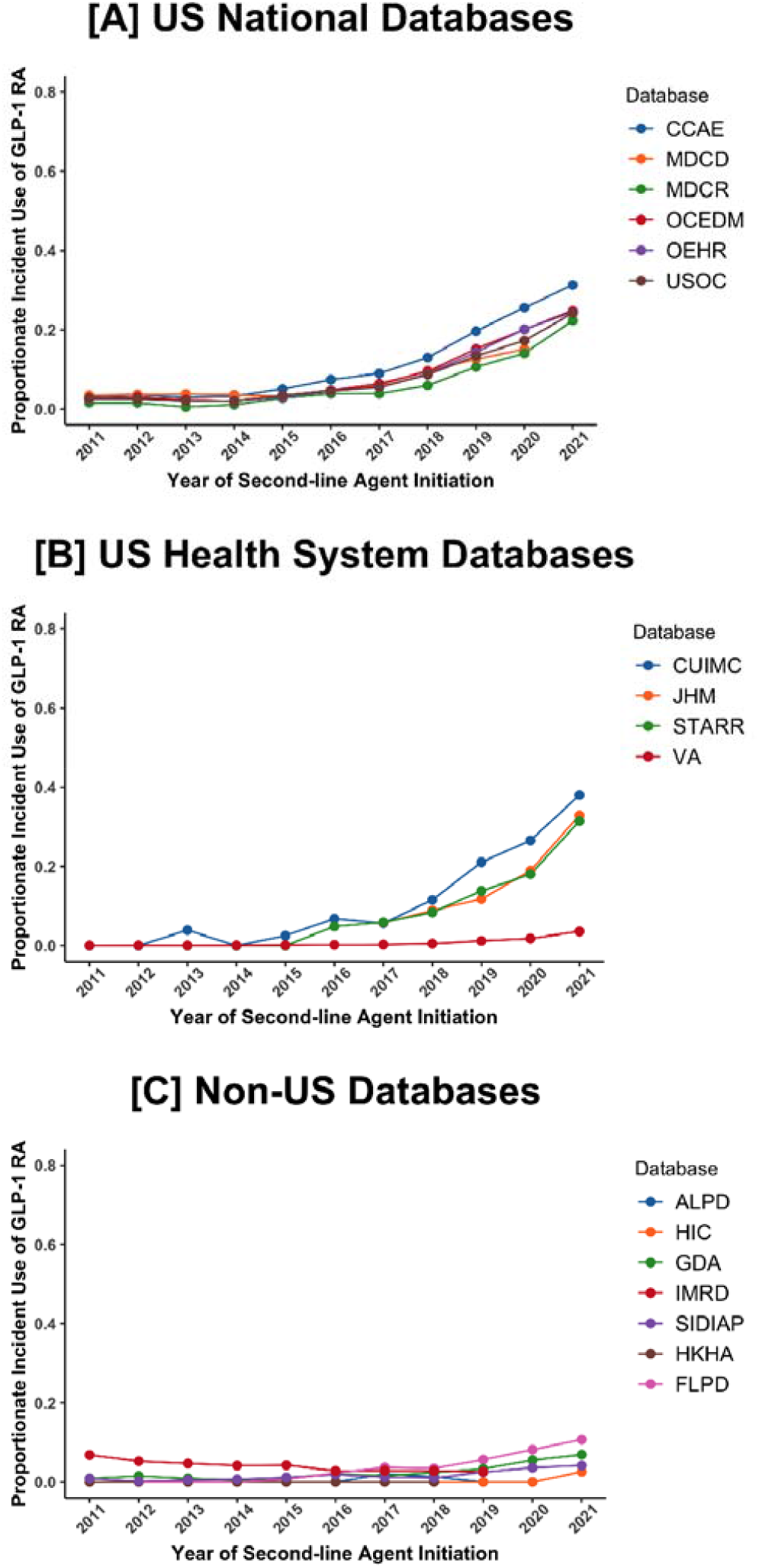
Proportional First Incident Use of Sodium-Glucose Cotransporter 2 Inhibitors as Second-line Therapy after Metformin in Patients with Established Cardiovascular Disease. **Abbreviations:** ALPD – Australia Longitudinal Patient Database Practice Profile, CCAE – IBM MarketScan® Commercial Claims and Encounters Data (CCAE), CUIMC – Columbia University Irving Medical Center, FLPD – France Longitudinal Patient Database, GDA – Germany Disease Analyser, HIC – Health Informatics Centre at the University of Dundee, HKHA - Hong Kong Hospital Authority, IMRD – UK-IQVIA Medical Research Data, JHM – Johns Hopkins Medicine, MDCD – IBM Health MarketScan® Multi-State Medicaid Database, MDCR – IBM Health MarketScan® Medicare Supplemental and Coordination of Benefits Database, OCEDM – Optum Clinformatics Extended Data Mart – Date of Death (DOD), OEHR – Optum© de-identified Electronic Health Record Dataset, SGLT2i - Sodium-Glucose Cotransporter 2 Inhibitor, SIDIAP - Information System for Research in Primary Care, STARR - Stanford Medicine, USOC - United States Open Claims, VA - Department of Veterans Affairs Healthcare System

**Figure 7.**
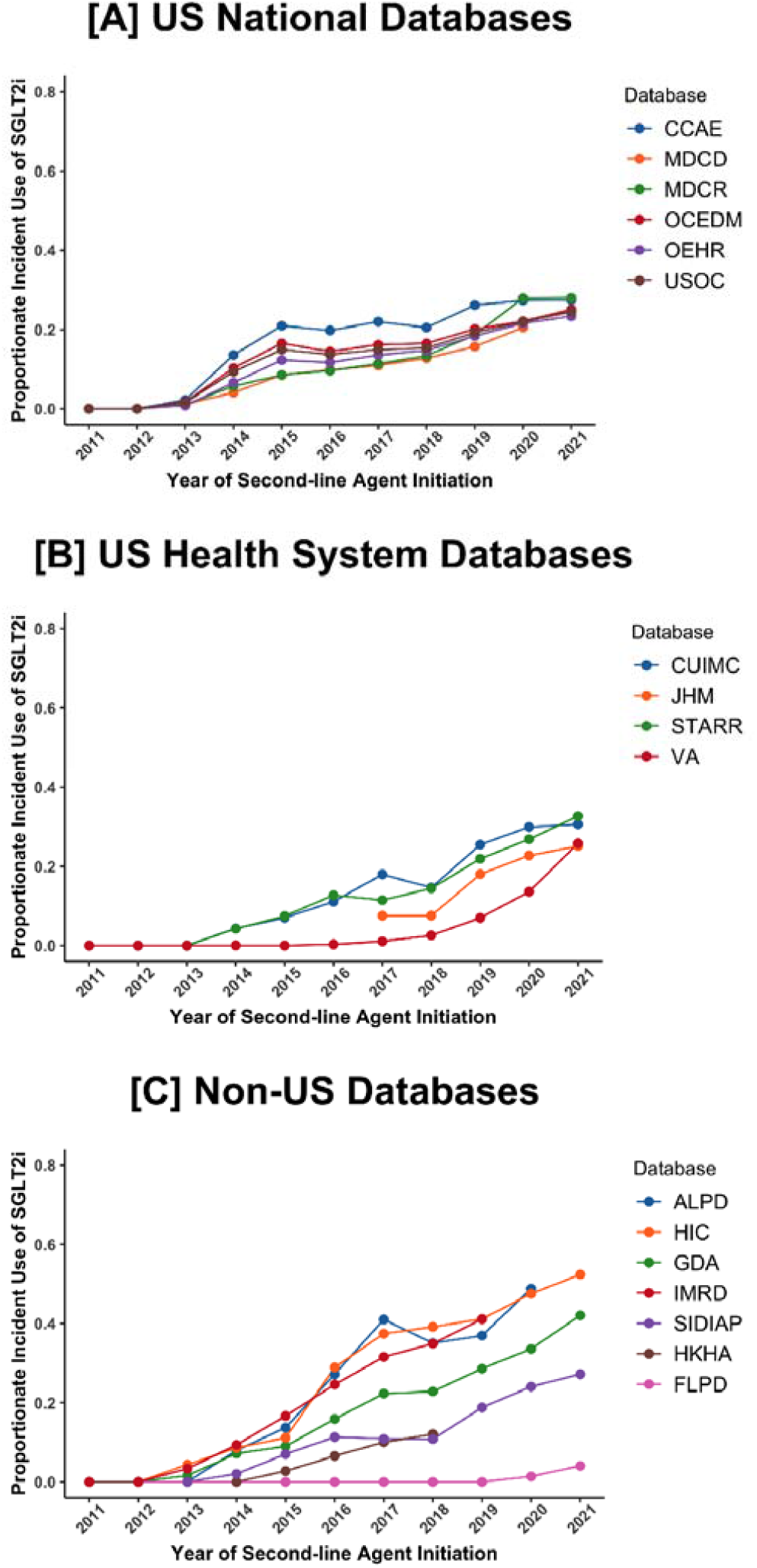
Proportional First Incident Use of Sodium-Glucose Cotransporter 2 Inhibitors as Second-line Therapy after Metformin in Patients without Established Cardiovascular Disease. **Abbreviations:** ALPD - Australia Longitudinal Patient Database Practice Profile, CCAE - IBM MarketScan® Commercial Claims and Encounters Data (CCAE), CUIMC - Columbia University Irving Medical Center, FLPD - France Longitudinal Patient Database, GDA - Germany Disease Analyser, HIC - Health Informatics Centre at the University of Dundee, HKHA - Hong Kong Hospital Authority, IMRD - UK-IQVIA Medical Research Data, JHM - Johns Hopkins Medicine, MDCD - IBM Health MarketScan® Multi-State Medicaid Database, MDCR - IBM Health MarketScan® Medicare Supplemental and Coordination of Benefits Database, OCEDM - Optum Clinformatics Extended Data Mart - Date of Death (DOD), OEHR - Optum© de-identified Electronic Health Record Dataset, SGLT2i - Sodium-Glucose Cotransporter 2 Inhibitor, SIDIAP - Information System for Research in Primary Care, STARR - Stanford Medicine, USOC – United States Open Claims, VA - Department of Veterans Affairs Healthcare System

Among the non-US health systems, the uptake of GLP-1 RAs increased from no uptake in 2011 to 13.4% in 2021 in patients with CVD in France, and to 10.7% in patients without CVD (**Figures 4 and 5**). While SGLT2is were not in use as second-line antihyperglycemic agents in 2011 in any of the non-US databases, their uptake grew to include 6.1% of the patients with CVD in France, and 54.2% of the patients with CVD in Scotland (**Figure 6**). In the patients without established CVD, the uptake of SGLT2is increased from no uptake in 2011 to 4.1% in France, and up to 52.3% in Australia in 2021 (**Figure 7**).

From 2016 to 2021, the uptake of GLP-1 RAs increased more significantly among patients without CVD compared to patients with CVD in France, UK, and some US databases; however, no database demonstrated higher annual change of GLP-1 RAs uptake in patients with CVD compared to patients without CVD (**Table 2**). There was a similar scenario for SGLT2is. Although Australia, UK, Scotland, and some US databases showed greater increases in the uptake of SGLT2is among patients without CVD compared to patients with CVD from 2016 to 2021, trends of the uptake of SGLT2is were not different between these populations in other databases (**Table 3**). The uptake trends of DPP-4is and SUs were inconsistent (**Supplemental Tables S12 and S13**).

**Table 2.**
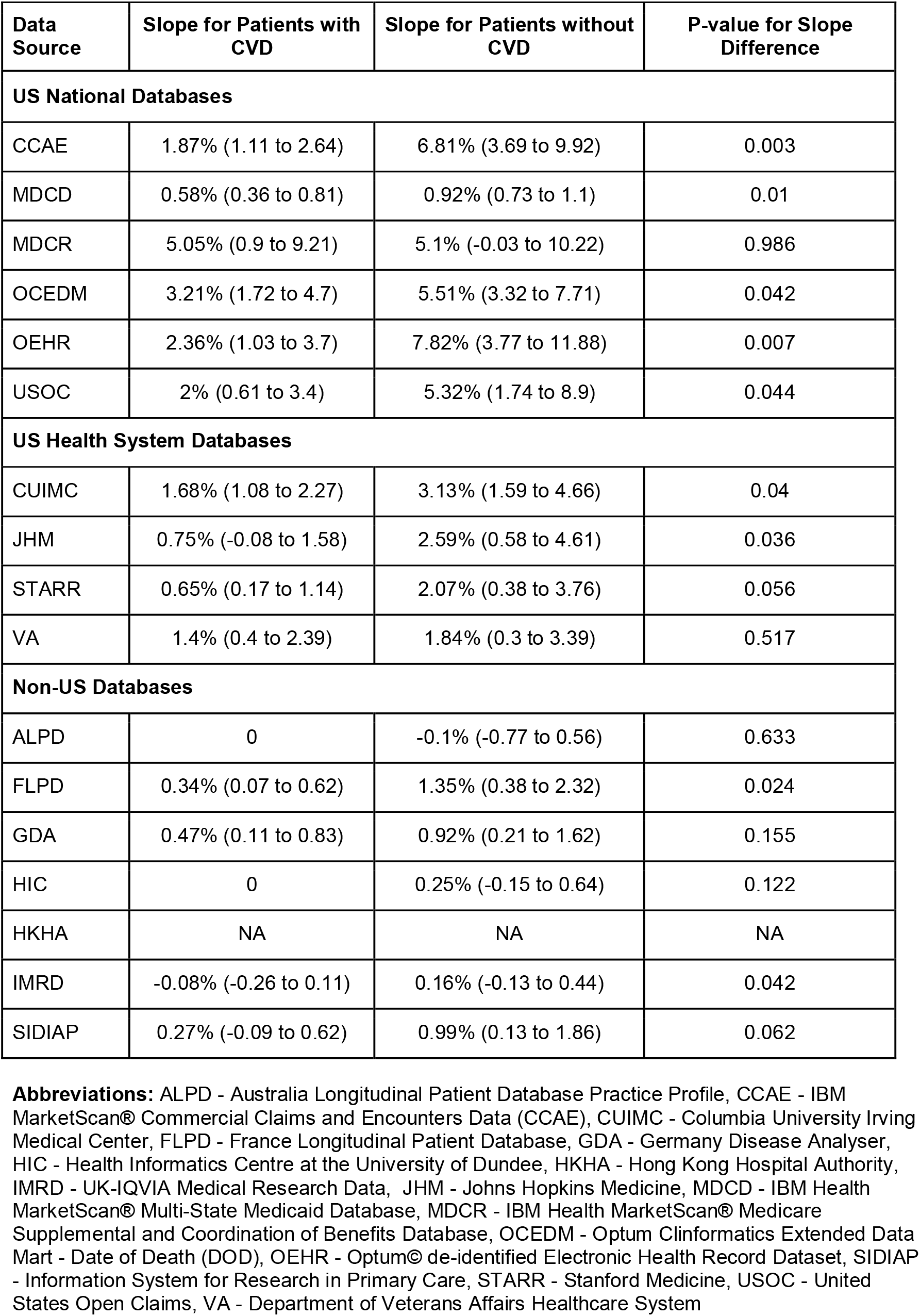
Annualized Change in the Incident Use of Glucagon-like Peptide-1 Receptor Agonists for Patients with Established Cardiovascular Disease and Patients without Established Cardiovascular Disease.

**Table 3.**
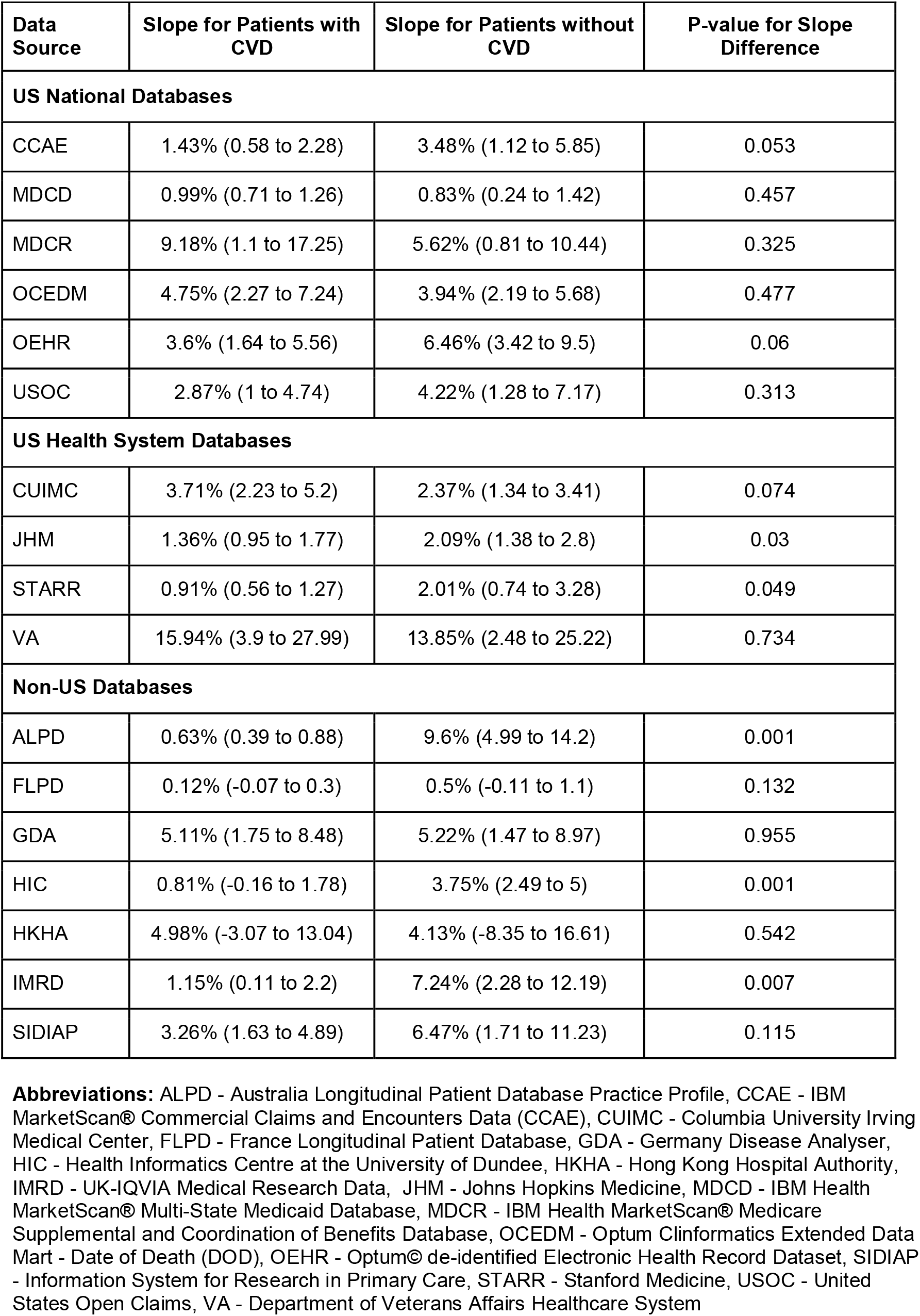
Annualized Change in the Incident Use of Sodium-Glucose Cotransporter 2 Inhibitors for Patients with Established Cardiovascular Disease and Patients without Established Cardiovascular Disease.

## DISCUSSION

In this first investigation from the LEGEND-T2DM study, we report a large and comprehensive pharmacoepidemiologic evaluation of the uptake of second-line T2DM agents across 17 international databases with over 4.8 million T2DM patient records. The study uses a federated approach to the study of patterns of medication use across multiple disparate data sources simultaneously, thereby allowing an informed assessment of individual trends in second-line T2DM medication uptake. We observed a large uptake of cardioprotective antihyperglycemic agents among patients initiating a second-line agent, representing nearly half of all included patients. While there was an increase in both cardioprotective drug classes in the US, the initiation of SGLT2is increased at a much higher rate, representing nearly a third of patients. In contrast, the initiation of SGLT2is increased to 40% to 50% of the population in a majority of European and Asian cohorts, with lower initiation of GLP-1 RAs. Finally, there are patterns suggestive of non-selective uptake of cardioprotective agents with an increasing uptake among those without CVD compared to those with established CVD, despite the latter group representing the only group with strong recommendations for use in clinical practice guidelines.[33,34]

The study builds upon previous assessments of GLP-1 RAs and SGLT2is use in both national US surveys and insurance datasets. These prior studies focused on the overall use of cardioprotective therapy and found that at most 10%-15% individuals with compelling indications use cardioprotective medications. [7,11,12,35,36] The current study adds to the literature by focusing on new initiators of second-line therapy, therefore, assessing initiation of these agents exclusively in individuals who likely required clinical escalation of antihyperglycemic therapy. Moreover, it represents the first assessment contrasting the trends observed in the US with those in other countries and demonstrating the large uptake of SGLT2is that has occurred in many countries in Europe and Asia, during a period when the use has been relatively limited in the US. We also find that the increase in GLP-1 RA initiation has been differential, with US patterns of measurable increase in GLP-1 RAs not translating in other countries.

The findings also suggest potential mechanisms for the patterns noted in the US. Initial studies finding low uptake for cardioprotective agents in the US had posited that this use may represent clinician inertia,[8] despite strong support in guidelines,[6,37] given the novel nature of these medications. This was suggested by the low use even among those with medical insurance. However, the rapid uptake in most nations with a nationally funded healthcare program with preventive medical coverage highlights that the underuse in the US may be financially motivated. While not evaluated in this study, these may include barriers associated with high out-of-pocket costs or other insurer-driven strategies to restrict their use.[38,39] This is particularly concerning in the US given the lack of requirement for commercial insurance to cover preventative therapy, for which a return on investment for insurers is often delayed.

A key exception to this pattern was France, where despite a national health insurance with prescription coverage,[40] the relative uptake of cardioprotective therapies was low. A review of clinical directives and guidelines in France suggests that national policies that urged caution against possible adverse events with novel agents may underlie these patterns.[41,42] The limited uptake of GLP-1 RAs in non-US despite their cardioprotective effects, may, however, indicate a barrier with their injectable nature, and the alternative available in SGLT2is with broader tolerability.[13,35,43] Therefore, financial rather than informational strategies are likely essential to promote the uptake of cardioprotective therapies in the US, particularly among those with CVD.

A key strength of our study is the demonstration of a novel strategy for monitoring medication use patterns on an international scale without the need for sharing of individual-level data, which can be easily adapted for monitoring the effect of local and international interventions. Our study has certain limitations. First, the study represents information from available datasets in respective countries and may not be all generalizable to the entire populations. Hence, they might not be all representative of the respective national populations. Second, there is potential for overlap of medical records in some US databases, such that the same patients may be captured across multiple sources. Having different record views of the same patient can be an advantage in capturing the real-life health events experienced by the patient. But, since licensing agreements prohibit attempts to link patients between most databases, the extent of this overlap cannot be precisely assessed. Third, given the heterogeneity in the included databases, it may be essential to standardize the patients on the basis of outcomes and assess the incident drug use. Fourth, although the drugs included are commonly used as second-line escalation therapy for T2DM, the precise reasons for initiation cannot be determined. Other potential reasons for prescription may include indications for weight loss, selection based on low cost of one agent over the other, or safer side-effect profile.

## CONCLUSION

Despite the increase in overall uptake of cardioprotective antihyperglycemic agents as second-line treatment for T2DM, there has been underuse of these agents in the US relative to other countries, particularly among those with established CVD. A strategy to ensure medication uptake concordant with guideline recommendations is essential to improve outcomes of patients with T2DM.

## Data Availability

The summary data can be accessed online at https://data.ohdsi.org/LegendT2dmClassCohortExplorer/ to allow exploration of the cohorts included in LEGEND-T2DM. Some of the datasets used within this study are available via license. The data that support the findings of this study are available to license from IBM (CCAE, MDCD, MDCR), Optum (OCEDM, OEHR), and IQVIA (IMRD). Data are available from IBM at https://www.ibm.com/products/marketscan-research-databases, from Optum at https://www.optum.com/business/solutions/life-sciences/real-world-data.html, and from IQVIA at https://www.iqvia.com/solutions/real-world-evidence/real-world-data-and-insights. Outside the license data previously described, this study was performed as a federated network study, meaning the data remained with the data partner. Individual organizations would need to be contacted in order to gain access to those data assets.

https://www.ibm.com/products/marketscan-research-databases

https://www.optum.com/business/solutions/life-sciences/real-world-data.html

https://www.iqvia.com/solutions/real-world-evidence/real-world-data-and-insights

## CONTRIBUTORS

All authors acquired, analyzed, and interpreted the data and critically revised the manuscript for important intellectual content. RK and MAS conceived and designed the study. RK, LSD, KL, JZ, AA, and MAS conducted the statistical analysis and drafted the manuscript. RK and MAS provided supervision. MAS is the study guarantor. RK and MAS had full access to all of the data in the study and take responsibility for the integrity of the data and the accuracy of the data analysis. The corresponding author attests that all listed authors meet authorship criteria and that no others meeting the criteria have been omitted.

## STATEMENTS

### Ethics Approval

All data sources received institutional review board approval or exemption for their participation in LEGEND-T2DM.

### Transparency Statement

The corresponding author affirms that the manuscript is an honest, accurate, and transparent account of the study being reported; that no important aspects of the study have been omitted; and that any discrepancies from the study as originally planned (and, if relevant, registered) have been explained.

### Dissemination to Participants and Related Patient and Public Communities

We will disseminate the results of the study through press release and social media postings to explain the result to news media and public.

### Copyright/License for Publication

The Corresponding Author has the right to grant on behalf of all authors and does grant on behalf of all authors, a worldwide licence to the Publishers and its licensees in perpetuity, in all forms, formats and media (whether known now or created in the future), to i) publish, reproduce, distribute, display and store the Contribution, ii) translate the Contribution into other languages, create adaptations, reprints, include within collections and create summaries, extracts and/or, abstracts of the Contribution, iii) create any other derivative work(s) based on the Contribution, iv) to exploit all subsidiary rights in the Contribution, v) the inclusion of electronic links from the Contribution to third party material where-ever it may be located; and, vi) licence any third party to do any or all of the above.

## SUMMARY BOX

### What is Already Known on this Topic

Cardioprotective second-line antihyperglycemic agents, glucagon-like peptide-1 receptor agonists and sodium-glucose cotransporter 2 inhibitors, have strong evidence not only of addressing hyperglycemia but also of improving cardiovascular risk in high-risk individuals with type 2 diabetes mellitus (T2DM). However, the actual uptake of these drugs continues to lag. Further, studies characterizing patterns of use have exclusively focused on prevalent use, and US-based studies have focused on single-payers or small populations included in national surveys.

### What this Study Adds

There was a large uptake in the use of cardioprotective antihyperglycemic agents among patients with T2DM initiating a second-line agent, representing nearly half of all patients across US and non-US cohorts. There are patterns suggestive of non-selective use of cardioprotective agents with an increasing uptake among those without cardiovascular disease compared with those with established atherosclerotic disease, despite the latter group representing the only group with strong recommendation for use in clinical practice guidelines.

