## Supplementary material for "Multinational Patterns of Second-line Anti-hyperglycemic Drug Initiation Across Cardiovascular Risk Groups: A Federated Pharmacoepidemiologic Evaluation in LEGEND-T2DM": Online Supplement

**Supplemental Methods – Exposure Cohort Definitions**

### I. Class-vs-Class Exposure (DPP4 New-User) Cohort / OT1

i. **Cohort Entry Events**

People with continuous observation of 365 days before the event may enter the cohort when observing any of the following:

1. drug exposure of ‘DPP4 inhibitors’ for the first time in the person’s history. Limit cohort entry events to the earliest event per person.

Restrict entry events to with all of the following criteria:

1. with the following event criteria: who are >= 18 years old.
2. having at least 1 condition occurrence of ‘Type 2 diabetes mellitus,’ starting anytime on or before cohort entry start date; allow events outside observation period.
3. having no condition occurrences of ‘Type 1 diabetes mellitus,’ starting anytime on or before cohort entry start date; allow events outside observation period.
4. having no condition occurrences of ‘Secondary diabetes mellitus,’ starting anytime on or before cohort entry start date; allow events outside observation period.
   ii. **Additional Inclusion Criteria**
       1. No prior GLP-1 receptor agonist exposure

Entry events having no drug exposures of ‘GLP-1 receptor agonists,’ starting anytime on or before cohort entry start date; allow events outside observation period.

2. No prior SGLT-2 inhibitor exposure

Entry events having no drug exposures of ‘SGLT2 inhibitors,’ starting anytime on or before cohort entry start date; allow events outside observation period.

3. No prior SU exposure

Entry events having no drug exposures of ‘Sulfonylureas,’ starting anytime on or before cohort entry start date; allow events outside observation period.

4. No prior other anti-diabetic exposure

Entry events having no drug exposures of ‘Other anti-diabetics,’ starting anytime on or before cohort entry start date; allow events outside observation period.

5. Prior metformin use

Entry events with any of the following criteria:

1. having at least 1 drug era of ‘Metformin,’ starting anytime up to 90 days before cohort entry start date; allow events outside observation period; with era length >= 90 days.
2. having at least 3 drug exposures of ‘Metformin,’ starting anytime on or before cohort entry start date; allow events outside observation period.
   - No prior insulin use or combo initiation: Proxy for < 30 days drug era anytime before index and no combination use on index

Entry events with all of the following criteria:

1. having no drug eras of ‘Insulin,’ starting anytime up to 30 days before cohort entry start date; allow events outside observation period; with era length > 30 days.
2. having no drug eras of ‘Insulin,’ starting between 30 days before and 0 days after cohort entry start date; allow events outside observation period.
   iii. **Cohort Exit**

The cohort end date will be based on a continuous exposure to ‘DPP4 inhibitors’: allowing 30 days between exposures, adding 0 days after exposure ends, and using days supply and exposure end date for exposure duration.

iv. **Cohort Eras**

Entry events will be combined into cohort eras if they are within 0 days of each other.

v. **Concept: DPP4 inhibitors**

| Concept ID | Concept Name | Code | Vocabulary | Excluded | Descendants | Mapped |
| --- | --- | --- | --- | --- | --- | --- |
| 43013884 | alogliptin | 1368001 | RxNorm | NO | YES | NO |
| 40239216 | linagliptin | 1100699 | RxNorm | NO | YES | NO |
| 40166035 | saxagliptin | 857974 | RxNorm | NO | YES | NO |
| 1580747 | sitagliptin | 593411 | RxNorm | NO | YES | NO |
| 19122137 | vildagliptin | 596554 | RxNorm | NO | YES | NO |

vi. **Concept: GLP-1 receptor agonists**

| Concept ID | Concept Name | Code | Vocabulary | Excluded | Descendants | Mapped |
| --- | --- | --- | --- | --- | --- | --- |
| 44816332 | albiglutide | 1534763 | RxNorm | NO | YES | NO |
| 45774435 | dulaglutide | 1551291 | RxNorm | NO | YES | NO |
| 1583722 | exenatide | 60548 | RxNorm | NO | YES | NO |
| 40170911 | liraglutide | 475968 | RxNorm | NO | YES | NO |
| 44506754 | lixisenatide | 1440051 | RxNorm | NO | YES | NO |
| 793143 | semaglutide | 1991302 | RxNorm | NO | YES | NO |

| Concept ID | Concept Name | Code | Vocabulary | Excluded | Descendants | Mapped |
| --- | --- | --- | --- | --- | --- | --- |
| 43526465 | canagliflozin | 1373458 | RxNorm | NO | YES | NO |
| 44785829 | dapagliflozin | 1488564 | RxNorm | NO | YES | NO |
| 45774751 | empagliflozin | 1545653 | RxNorm | NO | YES | NO |
| 793293 | ertugliflozin | 1992672 | RxNorm | NO | YES | NO |

vii. **Concept: SGLT2 inhibi****tors**
viii. **Concept: Sulfonylureas**

| Concept ID | Concept Name | Code | Vocabulary | Excluded | Descendants | Mapped |
| --- | --- | --- | --- | --- | --- | --- |
| 1594973 | chlorpropamide | 2404 | RxNorm | NO | YES | NO |
| 1597756 | glimepiride | 25789 | RxNorm | NO | YES | NO |
| 1560171 | glipizide | 4821 | RxNorm | NO | YES | NO |
| 19097821 | gliquidone | 25793 | RxNorm | NO | YES | NO |
| 1559684 | glyburide | 4815 | RxNorm | NO | YES | NO |
| 1502809 | tolazamide | 10633 | RxNorm | NO | YES | NO |
| 1502855 | tolbutamide | 10635 | RxNorm | NO | YES | NO |

ix. **Concept: Other anti-diabetics**

| Concept ID | Concept Name | Code | Vocabulary | Excluded | Descendants | Mapped |
| --- | --- | --- | --- | --- | --- | --- |
| 1529331 | acarbose | 16681 | RxNorm | NO | YES | NO |
| 1530014 | acetohexamide | 173 | RxNorm | NO | YES | NO |
| 730548 | bromocriptine | 1760 | RxNorm | NO | YES | NO |
| 19033498 | carbutamide | 2068 | RxNorm | NO | YES | NO |
| 19001409 | glibornuride | 102846 | RxNorm | NO | YES | NO |
| 19059796 | gliclazide | 4816 | RxNorm | NO | YES | NO |
| 19001441 | glymidine | 102848 | RxNorm | NO | YES | NO |
| 1510202 | miglitol | 30009 | RxNorm | NO | YES | NO |
| 1502826 | nateglinide | 274332 | RxNorm | NO | YES | NO |
| 1525215 | pioglitazone | 33738 | RxNorm | NO | YES | NO |
| 1516766 | repaglinide | 73044 | RxNorm | NO | YES | NO |
| 1547504 | rosiglitazone | 84108 | RxNorm | NO | YES | NO |
| 1515249 | troglitazone | 72610 | RxNorm | NO | YES | NO |

x. **Concept: Insulin**

| Concept ID | Concept Name | Code | Vocabulary | Excluded | Descendants | Mapped |
| --- | --- | --- | --- | --- | --- | --- |
| 1596977 | insulin, regular, human | 253182 | RxNorm | NO | YES | NO |
| 1550023 | insulin lispro | 86009 | RxNorm | NO | YES | NO |
| 1567198 | insulin aspart, human | 51428 | RxNorm | NO | YES | NO |
| 1502905 | insulin glargine | 274783 | RxNorm | NO | YES | NO |
| 1513876 | insulin lispro protamine, human | 314684 | RxNorm | NO | YES | NO |
| 1531601 | insulin aspart protamine, human | 352385 | RxNorm | NO | YES | NO |
| 1586346 | insulin, regular, pork | 221109 | RxNorm | NO | YES | NO |
| 1544838 | insulin glulisine, human | 400008 | RxNorm | NO | YES | NO |
| 1516976 | insulin detemir | 139825 | RxNorm | NO | YES | NO |
| 1590165 | insulin, regular, beef-pork | 235275 | RxNorm | NO | YES | NO |
| 1513849 | lente insulin, human | 314683 | RxNorm | NO | YES | NO |
| 1562586 | lente insulin, pork | 93108 | RxNorm | NO | YES | NO |
| 1588986 | insulin human, rDNA origin | 631657 | RxNorm | NO | YES | NO |
| 1513843 | lente insulin, beef-pork | 314682 | RxNorm | NO | YES | NO |
| 1586369 | ultralente insulin, human | 221110 | RxNorm | NO | YES | NO |
| 35605670 | insulin argine | 1740938 | RxNorm | NO | YES | NO |
| 35602717 | insulin degludec | 1670007 | RxNorm | NO | YES | NO |
| 21600713 | INSULINS AND ANALOGUES | A10A | ATC | NO | YES | NO |
| 19078608 | insulin, protamine zinc, beef-pork 100  UNT/ML Injectable Suspension | 311053 | RxNorm | NO | YES | NO |

xi. **Concept: Metformin**

| Concept ID | Concept Name | Code | Vocabulary | Excluded | Descendants | Mapped |
| --- | --- | --- | --- | --- | --- | --- |
| 1503297 | metformin | 6809 | RxNorm | NO | YES | NO |

xii. **Concept: Secondary diabetes mellitus**

| Concept ID Concept Name | Code | Vocabulary | Excluded | Descendants | Mapped |
| --- | --- | --- | --- | --- | --- |
| 195771 Secondary diabetes mellitus | 8801005 | SNOMED | NO | YES | NO |

xiii. **Concept: Type 1 diabetes mellitus**

| Concept ID | Concept Name | Code | Vocabulary | Excluded | Descendants | Mapped |
| --- | --- | --- | --- | --- | --- | --- |
| 201254 | Type 1 diabetes mellitus | 46635009 | SNOMED | NO | YES | NO |
| 435216 | Disorder due to type 1 diabetes mellitus | 420868002 | SNOMED | NO | YES | NO |
| 200687 | Renal disorder due to type 1 diabetes  mellitus | 421893009 | SNOMED | NO | YES | NO |
| 377821 | Disorder of nervous system due to type 1  diabetes mellitus | 421468001 | SNOMED | NO | YES | NO |
| 318712 | Peripheral circulatory disorder due to type 1 diabetes mellitus | 421365002 | SNOMED | NO | YES | NO |

xiv. **Concept: Type 2 diabetes mellitus**

| Concept ID | Concept Name | Code | Vocabulary | Excluded | Descendants | Mapped |
| --- | --- | --- | --- | --- | --- | --- |
| 201826 | Type 2 diabetes mellitus | 44054006 | SNOMED | NO | YES | NO |
| 443734 | Ketoacidosis due to type 2 diabetes mellitus | 421750000 | SNOMED | NO | YES | NO |
| 443767 | Disorder of eye due to diabetes mellitus | 25093002 | SNOMED | NO | YES | NO |
| 192279 | Disorder of kidney due to diabetes mellitus | 127013003 | SNOMED | NO | YES | NO |
| 443735 | Coma due to diabetes mellitus | 420662003 | SNOMED | NO | YES | NO |
| 376065 | Disorder of nervous system due to type 2  diabetes mellitus | 421326000 | SNOMED | NO | YES | NO |
| 443729 | Peripheral circulatory disorder due to type 2  diabetes mellitus | 422166005 | SNOMED | NO | YES | NO |
| 443732 | Disorder due to type 2 diabetes mellitus | 422014003 | SNOMED | NO | YES | NO |

### II. Metformin Use Modifier

i. **No prior metformin use**

Entry events having no drug eras of ‘Metformin,’ starting anytime on or before cohort entry start date; allow events outside observation period.

### III. Drug-vs-Drug Exposure (Alogliptin New-User) Cohort / OT1

i. **Cohort Entry Events**

People with continuous observation of 365 days before event may enter the cohort when observing any of the following:

1. drug exposure of ‘alogliptin’ for the first time in the person’s history. Limit cohort entry events to the earliest event per person.

Restrict entry events to with all of the following criteria:

1. with the following event criteria: who are >= 18 years old.
2. having at least 1 condition occurrence of ‘Type 2 diabetes mellitus,’ starting anytime on or before cohort entry start date; allow events outside observation period.
3. having no condition occurrences of ‘Type 1 diabetes mellitus,’ starting anytime on or before cohort entry start date; allow events outside observation period.
4. having no condition occurrences of ‘Secondary diabetes mellitus,’ starting anytime on or before cohort entry start date; allow events outside observation period.
   ii. **Additional Inclusion Criteria**

- No prior with-in class exposure

Entry events having no drug exposures of ‘DPP4 inhibitors excluding alogliptin,’ starting anytime on or before cohort entry start date; allow events outside observation period.

- No prior GLP-1 receptor agonist exposure

Entry events having no drug exposures of ‘GLP-1 receptor agonists,’ starting anytime on or before cohort entry start date; allow events outside observation period.

- No prior SGLT-2 inhibitor exposure

Entry events having no drug exposures of ‘SGLT2 inhibitors,’ starting anytime on or before cohort entry start date; allow events outside observation period.

- No prior SU exposure

Entry events having no drug exposures of ‘Sulfonylureas,’ starting anytime on or before cohort entry start date; allow events outside observation period.

- No prior other anti-diabetic exposure

Entry events having no drug exposures of ‘Other anti-diabetics,’ starting anytime on or before cohort entry start date; allow events outside observation period.

- Prior metformin use

Entry events with any of the following criteria:

1. having at least 1 drug era of ‘Metformin,’ starting anytime up to 90 days before cohort entry start date; allow events outside observation period; with era length >= 90 days.
2. having at least 3 drug exposures of ‘Metformin,’ starting anytime on or before cohort entry start date; allow events outside observation period.
   - No prior insulin use or combo initiation: Proxy for < 30 days drug era anytime before index and no combination use on index

Entry events having no drug eras of ‘Insulin,’ starting anytime on or before cohort entry start date; allow events outside observation period; with era length > 30 days.

iii. **Cohort Exit**

The cohort end date will be based on a continuous exposure to ‘alogliptin’: allowing 30 days between exposures, adding 0 days after exposure ends, and using days supply and exposure end date for exposure duration.

iv. **Cohort Eras**

Entry events will be combined into cohort eras if they are within 0 days of each other.

v. **Concept: alogliptin**

| Concept ID | Concept Name | Code Vocabulary | Excluded | Descendants | Mapped |
| --- | --- | --- | --- | --- | --- |
| 43013884 | alogliptin | 1368001 RxNorm | NO | YES | NO |

vi. **Concept: DPP4 inhibitors excluding alogliptin**

| Concept ID | Concept Name | Code | Vocabulary | Excluded | Descendants | Mapped |
| --- | --- | --- | --- | --- | --- | --- |
| 40239216 | linagliptin | 1100699 | RxNorm | NO | YES | NO |
| 40166035 | saxagliptin | 857974 | RxNorm | NO | YES | NO |
| 1580747 | sitagliptin | 593411 | RxNorm | NO | YES | NO |
| 19122137 | vildagliptin | 596554 | RxNorm | NO | YES | NO |

### IV. Heterogenity Study Inclusion Criteria

i. **Lower age group**

Entry events with the following event criteria: who are < 45 years old.

ii. **Middle age group**

Entry events with all of the following criteria:

1. with the following event criteria: who are >= 45 years old.
2. with the following event criteria: who are < 65 years old.
iii. **Older age group**

Entry events with the following event criteria: who are >= 65 years old.

iv. **Female stratum**

Entry events with the following event criteria: who are female.

v. **Male stratum**

Entry events with the following event criteria: who are male.

vi. **Race stratum**

Entry events with the following event criteria: race is: “black or african american,” “black,” “african american,” “african,” “bahamian,” “barbadian,” “dominican,” “dominica islander,” “haitian,” “jamaican,” “tobagoan,” “trinidadian” or “west indian.”

vii. **Low cardiovascular risk**

Entry events with all of the following criteria:

1. having no condition occurrences of ‘Conditions indicating established cardiovascular disease,’ starting anytime on or before cohort entry start date; allow events outside observation period.
2. having no procedure occurrences of ‘Procedures indicating established cardiovas- cular disease,’ starting anytime on or before cohort entry start date; allow events outside observation period.
viii. **Higher cardiovascular risk**

Entry events with any of the following criteria:

1. having at least 1 condition occurrence of ‘Conditions indicating established cardio- vascular disease,’ starting anytime on or before cohort entry start date; allow events outside observation period.
2. having at least 1 procedure occurrence of ‘Procedures indicating established cardio- vascular disease,’ starting anytime on or before cohort entry start date; allow events outside observation period.
ix. **Concept: Conditions indicating established cardiovascular disease**

| Concept ID | Concept Name | Code | Vocabulary | Excluded | Descendants | Mapped |
| --- | --- | --- | --- | --- | --- | --- |
| 319844 | Acute ischemic heart disease | 413439005 | SNOMED | NO | YES | NO |
| 321318 | Angina pectoris | 194828000 | SNOMED | NO | YES | NO |
| 4124841 | Aortic bifurcation syndrome | 233972005 | SNOMED | YES | YES | NO |
| 312337 | Arterial embolus and thrombosis | 266262004 | SNOMED | NO | YES | NO |
| 4278217 | Arterial thrombosis | 65198009 | SNOMED | NO | YES | NO |
| 40484167 | Arteriosclerosis of artery of extremity | 443971004 | SNOMED | NO | YES | NO |
| 318443 | Arteriosclerotic vascular disease | 72092001 | SNOMED | NO | YES | NO |
| 314659 | Arteritis | 52089001 | SNOMED | NO | NO | NO |
| 40479625 | Atherosclerosis of artery | 441574008 | SNOMED | NO | YES | NO |
| 40484541 | Atherosclerosis of autologous vein bypass  graft of limb | 442693003 | SNOMED | YES | YES | NO |
| 312902 | Benign intracranial hypertension | 68267002 | SNOMED | YES | YES | NO |
| 4288310 | Carotid artery obstruction | 69798007 | SNOMED | YES | YES | NO |
| 372924 | Cerebral artery occlusion | 20059004 | SNOMED | NO | YES | NO |
| 376713 | Cerebral hemorrhage | 274100004 | SNOMED | NO | YES | NO |
| 381591 | Cerebrovascular disease | 62914000 | SNOMED | NO | YES | NO |
| 316494 | Cerebrovascular disorder in the puerperium | 6594005 | SNOMED | YES | YES | NO |
| 315286 | Chronic ischemic heart disease | 413838009 | SNOMED | NO | YES | NO |
| 44782819 | Chronic occlusion of artery of extremity | 698816006 | SNOMED | NO | YES | NO |
| 4313767 | Chronic peripheral venous hypertension | 423674003 | SNOMED | YES | YES | NO |
| 372721 | Congenital anomaly of cerebrovascular  system | 65587001 | SNOMED | YES | YES | NO |
| 316995 | Coronary occlusion | 63739005 | SNOMED | NO | YES | NO |
| 134057 | Disorder of cardiovascular system | 49601007 | SNOMED | NO | NO | NO |
| 40480453 | Disorder of vein of lower extremity | 441739009 | SNOMED | YES | YES | NO |
| 46272492 | Dissection of artery | 710864009 | SNOMED | YES | YES | NO |
| 4324690 | Fracture of skull | 71642004 | SNOMED | YES | YES | NO |
| 441246 | Hemangioma of intracranial structure | 93468003 | SNOMED | YES | YES | NO |
| 380113 | Hemorrhage in optic nerve sheaths | 14460007 | SNOMED | YES | YES | NO |
| 192763 | Injury of blood vessel | 57662003 | SNOMED | YES | YES | NO |
| 4275428 | Injury of vein | 64583005 | SNOMED | YES | YES | NO |
| 442774 | Intermittent claudication | 63491006 | SNOMED | NO | YES | NO |
| 439847 | Intracranial hemorrhage | 1386000 | SNOMED | NO | YES | NO |
| 434056 | Late effects of cerebrovascular disease | 195239002 | SNOMED | NO | YES | NO |
| 4146311 | Leriche’s syndrome | 307816004 | SNOMED | NO | YES | NO |
| 4329847 | Myocardial infarction | 22298006 | SNOMED | NO | YES | NO |
| 4296029 | Periarteritis | 76805007 | SNOMED | NO | YES | NO |
| 260841 | Perinatal subarachnoid hemorrhage | 21202004 | SNOMED | YES | YES | NO |
| 317309 | Peripheral arterial occlusive disease | 399957001 | SNOMED | NO | YES | NO |
| 321822 | Peripheral vascular disorder due to diabetes  mellitus | 421895002 | SNOMED | NO | YES | NO |
| 313928 | Peripheral vascular complication | 10596002 | SNOMED | NO | YES | NO |
| 321052 | Peripheral vascular disease | 400047006 | SNOMED | NO | NO | NO |
| 44782775 | Peripheral vascular disease associated with  another disorder | 34881000119105 | SNOMED | NO | YES | NO |
| 318137 | Phlebitis and thrombophlebitis of intracranial  sinuses | 192753009 | SNOMED | YES | YES | NO |
| 441039 | Phlebitis of lower limb vein | 312588002 | SNOMED | NO | YES | NO |
| 4067424 | Polyarteritis | 20258000 | SNOMED | NO | YES | NO |
| 320749 | Polyarteritis nodosa | 155441006 | SNOMED | YES | YES | NO |
| 443239 | Precerebral arterial occlusion | 266253001 | SNOMED | NO | YES | NO |
| 440417 | Pulmonary embolism | 59282003 | SNOMED | YES | YES | NO |
| 4318842 | Renal vasculitis | 95578000 | SNOMED | NO | YES | NO |
| 380943 | Rupture of syphilitic cerebral aneurysm | 186893003 | SNOMED | YES | YES | NO |
| 432923 | Subarachnoid hemorrhage | 21454007 | SNOMED | NO | YES | NO |
| 439040 | Subdural hemorrhage | 35486000 | SNOMED | NO | YES | NO |
| 320741 | Thrombophlebitis | 64156001 | SNOMED | YES | YES | NO |
| 4141106 | Thrombosis of arteries of the extremities | 33591000 | SNOMED | NO | YES | NO |
| 4132546 | Traumatic brain injury | 127295002 | SNOMED | YES | YES | NO |
| 4194610 | Trunk arterial embolus | 312593004 | SNOMED | NO | YES | NO |
| 318169 | Varicose veins of lower extremity | 72866009 | SNOMED | YES | YES | NO |
| 4189293 | Vascular disorder of lower extremity | 373408007 | SNOMED | NO | YES | NO |
| 443752 | Ventricular hemorrhage | 23276006 | SNOMED | YES | YES | NO |
| 432346 | Dissection of vertebral artery | 230730001 | SNOMED | YES | YES | NO |

x. **Concept: Procedures indicating established cardiovascular disease**

| Concept ID | Concept Name | Code | Vocabulary | Excluded | Descendants | Mapped |
| --- | --- | --- | --- | --- | --- | --- |
| 4150819 | Operative procedure on coronary artery | 31413008 | SNOMED | NO | YES | NO |
| 4331725 | Operative procedure on artery of extremity | 22701007 | SNOMED | NO | YES | NO |

xi. **Without renal impairment**

Entry events having no condition occurrences of ‘Renal impairment,’ starting anytime on or before cohort entry start date; allow events outside observation period.

xii. **Renal impairment**

Entry events having at least 1 condition occurrence of ‘Renal impairment,’ starting anytime on or before cohort entry start date; allow events outside observation period.

xiii. **Concept: Renal impairment**

Concept ID	Concept Name	Code	Vocabulary	Excluded	Descendants	Mapped

4030518	Renal impairment

236423003	SNOMED	NO

YES

NO

### V. Escalation Exit Criteria

The cohort end date will be based on a continuous exposure to ‘DPP4 inhibitors’: allowing 30 days between exposures, adding 0 days after exposure ends, and using days supply and exposure end date for exposure duration.

The person also exists the cohort when encountering any of the following events:

1. drug exposures of ‘All alternative target exposures.’
2. drug exposures of ‘Other anti-diabetics.’
3. drug eras of ‘Insulin,’ with era length > 30 days.
   i. **Concept: All alternative target exposures**

| Concept ID | Concept Name | Code | Vocabulary | Excluded | Descendants | Mapped |
| --- | --- | --- | --- | --- | --- | --- |
| 44816332 | albiglutide | 1534763 | RxNorm | NO | YES | NO |
| 43526465 | canagliflozin | 1373458 | RxNorm | NO | YES | NO |
| 1594973 | chlorpropamide | 2404 | RxNorm | NO | YES | NO |
| 44785829 | dapagliflozin | 1488564 | RxNorm | NO | YES | NO |
| 45774435 | dulaglutide | 1551291 | RxNorm | NO | YES | NO |
| 45774751 | empagliflozin | 1545653 | RxNorm | NO | YES | NO |
| 793293 | ertugliflozin | 1992672 | RxNorm | NO | YES | NO |
| 1583722 | exenatide | 60548 | RxNorm | NO | YES | NO |
| 1597756 | glimepiride | 25789 | RxNorm | NO | YES | NO |
| 1560171 | glipizide | 4821 | RxNorm | NO | YES | NO |
| 19097821 | gliquidone | 25793 | RxNorm | NO | YES | NO |
| 1559684 | glyburide | 4815 | RxNorm | NO | YES | NO |
| 40170911 | liraglutide | 475968 | RxNorm | NO | YES | NO |
| 44506754 | lixisenatide | 1440051 | RxNorm | NO | YES | NO |
| 793143 | semaglutide | 1991302 | RxNorm | NO | YES | NO |
| 1502809 | tolazamide | 10633 | RxNorm | NO | YES | NO |
| 1502855 | tolbutamide | 10635 | RxNorm | NO | YES | NO |

**Supplemental Table S1 |** **Brief Descriptions of Databases from the Observational Health Data Sciences and Informatics Network Included in the Study**

| **Name of Database** | **Abbreviation** | **Brief Description** |
| --- | --- | --- |
| **US National Databases** | | |
| IBM MarketScan® Commercial Claims and Encounters Data | CCAE | IBM Health MarketScan® Commercial Claims and Encounters Database (CCAE) represent data from individuals enrolled in United States employer-sponsored insurance health plans. The data includes adjudicated health insurance claims (e.g. inpatient, outpatient, and outpatient pharmacy) as well as enrollment data from large employers and health plans who provide private healthcare coverage to employees, their spouses, and dependents. Additionally, it captures laboratory tests for a subset of the covered lives. This administrative claims database includes a variety of fee-for-service, preferred provider organizations, and capitated health plans. |
| IBM Health MarketScan® Multi-State Medicaid Database | MDCD | IBM MarketScan® Multi-State Medicaid Database (MDCD) adjudicated US health insurance claims for Medicaid enrollees from multiple states and includes hospital discharge diagnoses, outpatient diagnoses and procedures, and outpatient pharmacy claims as well as ethnicity and Medicare eligibility. Members maintain their same identifier even if they leave the system for a brief period however the dataset lacks lab data. |
| IBM Health MarketScan Medicare Supplemental and Coordination of Benefits Database | MDCR | IBM Health MarketScan® Medicare Supplemental and Coordination of Benefits Database (MDCR) represents health services of retirees in the United States with primary or Medicare supplemental coverage through privately insured fee-for-service, point-of-service, or capitated health plans. These data include adjudicated health insurance claims (e.g. inpatient, outpatient, and outpatient pharmacy). Additionally, it captures laboratory tests for a subset of the covered lives. |
| Optum Clinformatics Extended Data Mart - Date of Death (DOD) | OCEDM | Optum Clinformatics Extended DataMart is an adjudicated US administrative health claims database for members of private health insurance, who are fully insured in commercial plans or in administrative services only (ASOs), Legacy Medicare Choice Lives (prior to January 2006), and Medicare Advantage (Medicare Advantage Prescription Drug coverage starting January 2006). The population is primarily representative of commercial claims patients (0-65 years old) with some Medicare (65+ years old) however ages are capped at 90 years. It includes data captured from administrative claims processed from inpatient and outpatient medical services and prescriptions as dispensed, as well as results for outpatient lab tests processed by large national lab vendors who participate in data exchange with Optum. This dataset also provides date of death (month and year only) for members with both medical and pharmacy coverage from the Social Security Death Master File (however after 2011 reporting frequency changed due to changes in reporting requirements) and location information for patients is at the US state level. |
| Optum© de-identified Electronic Health Record Dataset | OEHR | Optum© de-identified Electronic Health Record Dataset represents Humedica’s Electronic Health Record data a medical records database. The medical record data includes clinical information, inclusive of prescriptions as prescribed and administered, lab results, vital signs, body measurements, diagnoses, procedures, and information derived from clinical Notes using Natural Language Processing (NLP). |
| US Open Claims | USOC | US Open Claims is a United States database of open, pre-adjudicated claims from 2000 to present. Data are reported at anonymized patient level collected from office-based physicians and specialists via office management software and clearinghouse switch sources for the purpose of reimbursement. A subset of medical claims data has adjudicated claims. |
| **US Health System Databases** | | |
| Columbia University Irving Medical Center | CUIMC | The Columbia University Irving Medical Center (CUIMC) database comprises electronic health records on 6,666,613 patients, with data collection starting in 1985. CUIMC is a northeast US quaternary care center with primary care practices in northern Manhattan and surrounding areas, and the database includes inpatient and outpatient care. The database currently holds information about the person (demographics), visits (inpatient and outpatient), conditions (billing diagnoses and problem lists), drugs (outpatient prescriptions and inpatient orders and administrations), devices, measurements (laboratory tests and vital signs), and other observations (symptoms). The data sources include current and previous electronic health record systems (homegrown Clinical Information System, homegrown WebCIS, Allscripts Sunrise Clinical Manager, Allscripts TouchWorks, Epic Systems), administrative systems (IBM PCS-ADS, Eagle Registration, IDX Systems, Epic Systems), and ancillary systems (homegrown LIS, Sunquest, Cerner Laboratory). |
| Johns Hopkins Medicine | JHM | The Johns Hopkins Medicine (JHM) database comprises electronic health records on 2.58 million patients, with data collection starting in 2016. JHM is a northeast US quaternary care center with inpatient hospitals and outpatient care centers in Baltimore, Maryland and the surrounding Chesapeake area. |
| Stanford Medicine | STARR | STAnford medicine Research data Repository, a clinical data warehouse containing live Epic data from Stanford Health Care, the Stanford Children’s Hospital, the University Healthcare Alliance and Packard Children's Health Alliance clinics and other auxiliary data from Hospital applications such as radiology PACS. STARR platform is developed and operated by Stanford Medicine Research IT team and is made possible by Stanford School of Medicine Research Office.[44] |
| Department of Veterans Affairs health care system | VA | VA OMOP data reflects the national Department of Veterans Affairs health care system, which is the largest integrated provider of medical and mental health services in the United States. Care is provided at 170 VA Medical Centers and 1,063 outpatient sites serving more than 9 million enrolled Veterans each year. |
| **Non-US Databases** | | |
| Australia Longitudinal Patient Database and Practice Profile | ALPD | Australia Electronic Medical Record is comprised of anonymized patient records collected from patient management software used by general practitioners to document patients’ clinical records. Data are collected from 2 sources (LPD – Longitudinal Patient Data and PP – Practice Profiles). LPD and PP data comes through in different tables and is integrated into one common data source. This data coverages primary care and general practices mainly for office-based patients. Data coverage includes over 2.9M patient records with at least one visit. Dates of service include from 2012 through present. Observation time is defined by the first and last consultation dates. Drugs are captured as prescription records with product, quantity, dosing directions, strength, indication and date of consultation. |
| France Longitudinal Patient Database | FLPD | France Longitudinal Patient Database is a computerized network of physicians including general practitioners who contribute to a centralized database of anonymized patient EMR. The database covers a time period from 2012 through the present. Observation time is defined by the first and last consultation dates. Drug information is derived from GP prescriptions. Drugs obtained over the counter by the patient outside the prescription system are not reported. No explicit registration or approval is necessary for drug utilization studies. |
| Germany Disease Analyser | GDA | Germany Disease Analyser is collected from extracts of patient management software used by general practitioners and specialists practicing in ambulatory care settings. Data coverage includes 40.2M distinct person records, about 48.2% population in the country and collected from 2.8K providers. Patient visiting more than one provider are not cross identified for data protection reasons and therefore recorded as separate in the system. Dates of service include from 1992 through present. Observation time is defined by the first and last consultation dates. Germany has no mandatory general practitioner system and patient have free choice of specialist. Drugs are recorded as prescriptions of marketed products. No registration or approval is required for drug utilization studies. |
| Health Informatics Centre at the University of Dundee | HIC | Health datasets covering approximately 1.2M people from the Tayside and Fife regions of Scotland, provided by the Health Informatics Centre (HIC) at the University of Dundee. |
| Hong Kong Hospital Authority | HKHA | Hong Kong Hospital Authority is the only regulatory body for all public hospitals in Hong Kong, which include 43 hospitals and institutions, 49 specialist Out-patient Clinics, and 73 general Out-patient Clinics. The electronic health record contains data on patient demographics, prescriptions, and diagnoses with real-time updates for routine clinical management used. |
| Information System for Research in Primary Care | SIDIAP | The Information System for Research in Primary Care (SIDIAP; www.sidiap.org) is a primary care records database that covers approximately 80% of the population of Catalonia, North-East Spain.[45] Healthcare is universal and tax-payer funded in the region, and primary care physicians are gatekeepers for all care and responsible for repeat prescriptions. |
| UK-IQVIA Medical Research Data | IMRD | The UK-IQVIA Medical Research Data (IMRD), previously known as The Health Improvement Network (THIN), contains anonymized electronic health records from over 744 general practices in the UK, covering approximately 6% of the UK population. It contains data on prescriptions, diagnoses, referrals, and patient demographics broadly representative of the UK. |

**Supplemental Table S2 |** **Pharmacological Agents Included in the Drug Classes**

| **GLP-1 RA** | **SGLT2i** | **DPP-4i** | **SU** |
| --- | --- | --- | --- |
| Albiglutide | Canagliflozin | Alogliptin | Chlorpropamide |
| Dulaglutide | Dapagliflozin | Linagliptin | Glimepiride |
| Exenatide | Empagliflozin | Saxagliptin | Glipizide |
| Liraglutide | Ertugliflozin | Sitagliptin | Gliquidone |
| Lixisenatide |  | Vildagliptin | Glyburide |
| Semaglutide |  |  | Tolazamide |
|  |  |  | Tolbutamide |

**Abbreviations:** DPP-4i - Dipeptidyl Peptidase-4 Inhibitors, GLP-1 RA - Glucagon-like Peptide-1 Receptor Agonist, SGLT2i - Sodium-Glucose Cotransporter 2 Inhibitor, SU - Sulfonylurea

**Supplemental Table S3 | Number of Participants of Databases from the Observational Health Data Sciences and Informatics Network Included in the Study by Calendar Year**

| **Database** | **Cohort Counts by Calendar Year** | | | | | | | | | | |
| --- | --- | --- | --- | --- | --- | --- | --- | --- | --- | --- | --- |
|  | **2011** | **2012** | **2013** | **2014** | **2015** | **2016** | **2017** | **2018** | **2019** | **2020** | **2021** |
| **US National Databases** | | | | | | | | | | | |
| CCAE | 23507 | 27701 | 21648 | 26728 | 26621 | 27894 | 25953 | 23826 | 26827 | 26400 | 8769 |
| MDCD | 1671 | 1682 | 2241 | 2607 | 5315 | 6254 | 6540 | 5500 | 5353 | 2901 | NA |
| MDCR | 5597 | 5193 | 5279 | 5570 | 4616 | 4874 | 3528 | 2117 | 2478 | 2098 | 2507 |
| OCEDM | 12326 | 13095 | 13677 | 13987 | 15868 | 18032 | 23039 | 24939 | 27873 | 30043 | 18998 |
| OEHR | 11404 | 15230 | 21046 | 26266 | 34297 | 37556 | 38057 | 35986 | 38817 | 32346 | 8003 |
| USOC | 158201 | 202498 | 257454 | 301627 | 349550 | 364960 | 375617 | 372409 | 390281 | 410644 | 337950 |
| **US Health System Databases** | | | | | | | | | | | |
| CUIMC | 181 | 200 | 216 | 219 | 396 | 506 | 537 | 532 | 613 | 638 | 523 |
| JHM | NA | NA | NA | NA | NA | 0 | 211 | 669 | 794 | 841 | 1244 |
| STARR | 65 | 84 | 152 | 148 | 260 | 287 | 362 | 397 | 423 | 427 | 537 |
| VA | 19816 | 18579 | 18546 | 19247 | 20076 | 19619 | 21181 | 22395 | 24202 | 24319 | 22039 |
| **Non-US Databases** | | | | | | | | | | | |
| ALPD | 0 | 85 | 125 | 230 | 239 | 216 | 323 | 499 | 381 | 309 | 54 |
| FLPD | 0 | 470 | 1459 | 1377 | 1325 | 1308 | 1420 | 1504 | 1705 | 1604 | 1098 |
| GDA | 1561 | 2009 | 2114 | 2365 | 2712 | 3076 | 3885 | 4102 | 4437 | 4772 | 1409 |
| HIC | 364 | 354 | 360 | 369 | 433 | 522 | 606 | 593 | 639 | 609 | 731 |
| HKHA | 436 | 460 | 651 | 562 | 538 | 588 | 584 | 795 | NA | NA | NA |
| IMRD | 2335 | 2684 | 2691 | 2840 | 3401 | 3508 | 3557 | 3267 | 890 | NA | NA |
| SIDIAP | 4131 | 4457 | 4117 | 4191 | 4911 | 5735 | 6039 | 6681 | 8328 | 6861 | 5931 |

**Abbreviations:** ALPD - Australia Longitudinal Patient Database Practice Profile, CCAE - IBM MarketScan® Commercial Claims and Encounters Data (CCAE), CUIMC - Columbia University Irving Medical Center, FLPD - France Longitudinal Patient Database, GDA - Germany Disease Analyser, HIC - Health Informatics Centre at the University of Dundee, HKHA - Hong Kong Hospital Authority, IMRD - UK-IQVIA Medical Research Data, JHM - Johns Hopkins Medicine, MDCD - IBM Health MarketScan® Multi-State Medicaid Database, MDCR - IBM Health MarketScan® Medicare Supplemental and Coordination of Benefits Database, OCEDM - Optum Clinformatics Extended Data Mart - Date of Death (DOD), OEHR - Optum© de-identified Electronic Health Record Dataset, SIDIAP - Information System for Research in Primary Care, STARR - Stanford Medicine, USOC - United States Open Claims, VA - Department of Veterans Affairs Healthcare System

**Supplemental Table S4 |** **Baseline Characteristics and Clinical Covariates in Patients Using Glucagon-like Peptide-1 Receptor Agonists as Second-Line Antihyperglycemic Agents in Databases in the United States**

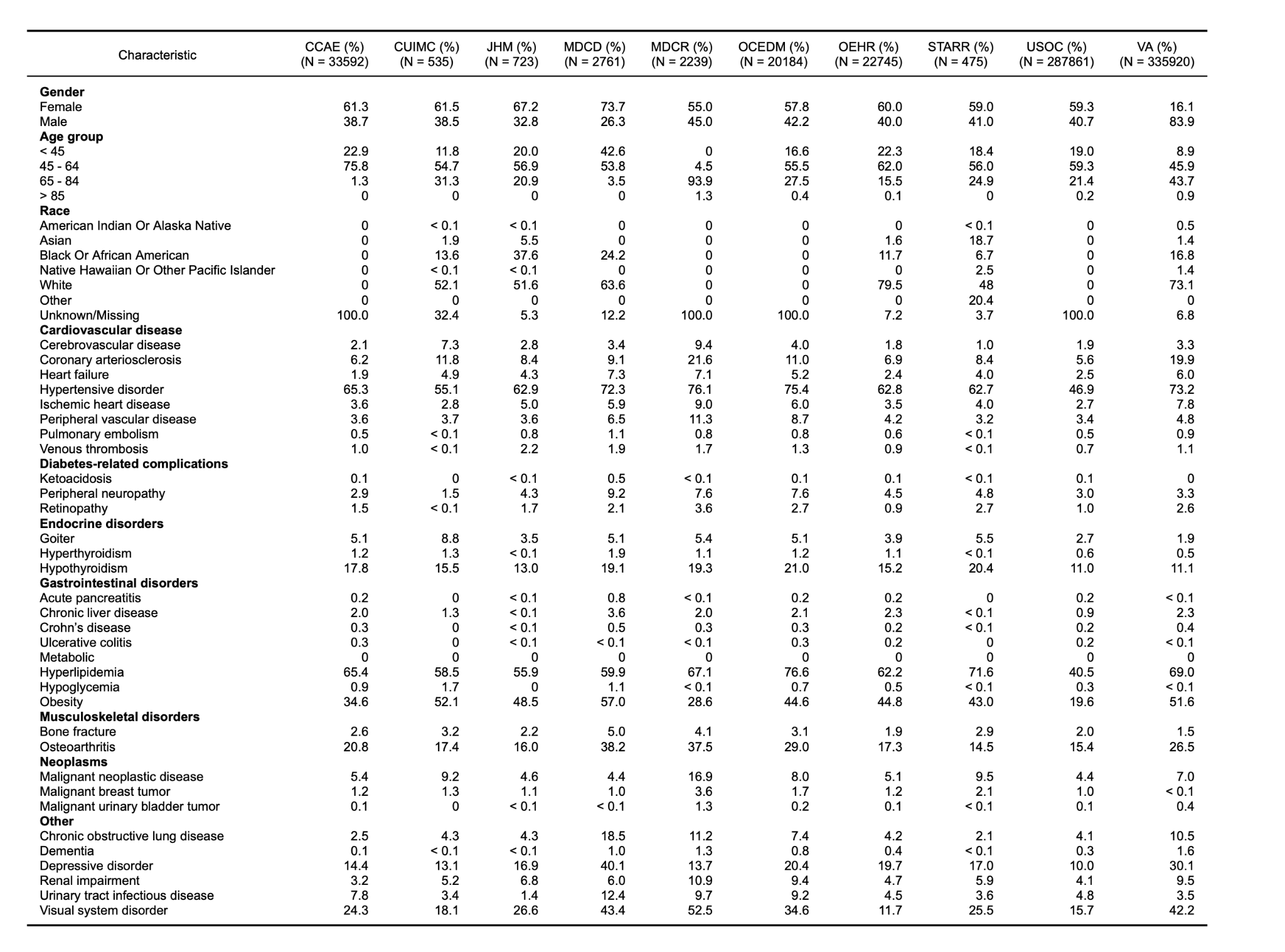

**Abbreviations:** CCAE: IBM MarketScan® Commercial Claims and Encounters Data (CCAE), CUIMC: Columbia University Irving Medical Center, JHM: Johns Hopkins Medicine, MDCD: IBM Health MarketScan® Multi-State Medicaid Database, MDCR: IBM Health MarketScan® Medicare Supplemental and Coordination of Benefits Database, OCEDM: Optum Clinformatics Extended Data Mart - Date of Death (DOD), OEHR: Optum© de-identified Electronic Health Record Dataset, STARR: Stanford Medicine, USOC: US Open Claims

**Supplemental Table S5 |** **Baseline Characteristics and Clinical Covariates in Patients Using Sodium-Glucose Cotransporter 2 Inhibitors as Second-Line Antihyperglycemic Agents in US Databases**

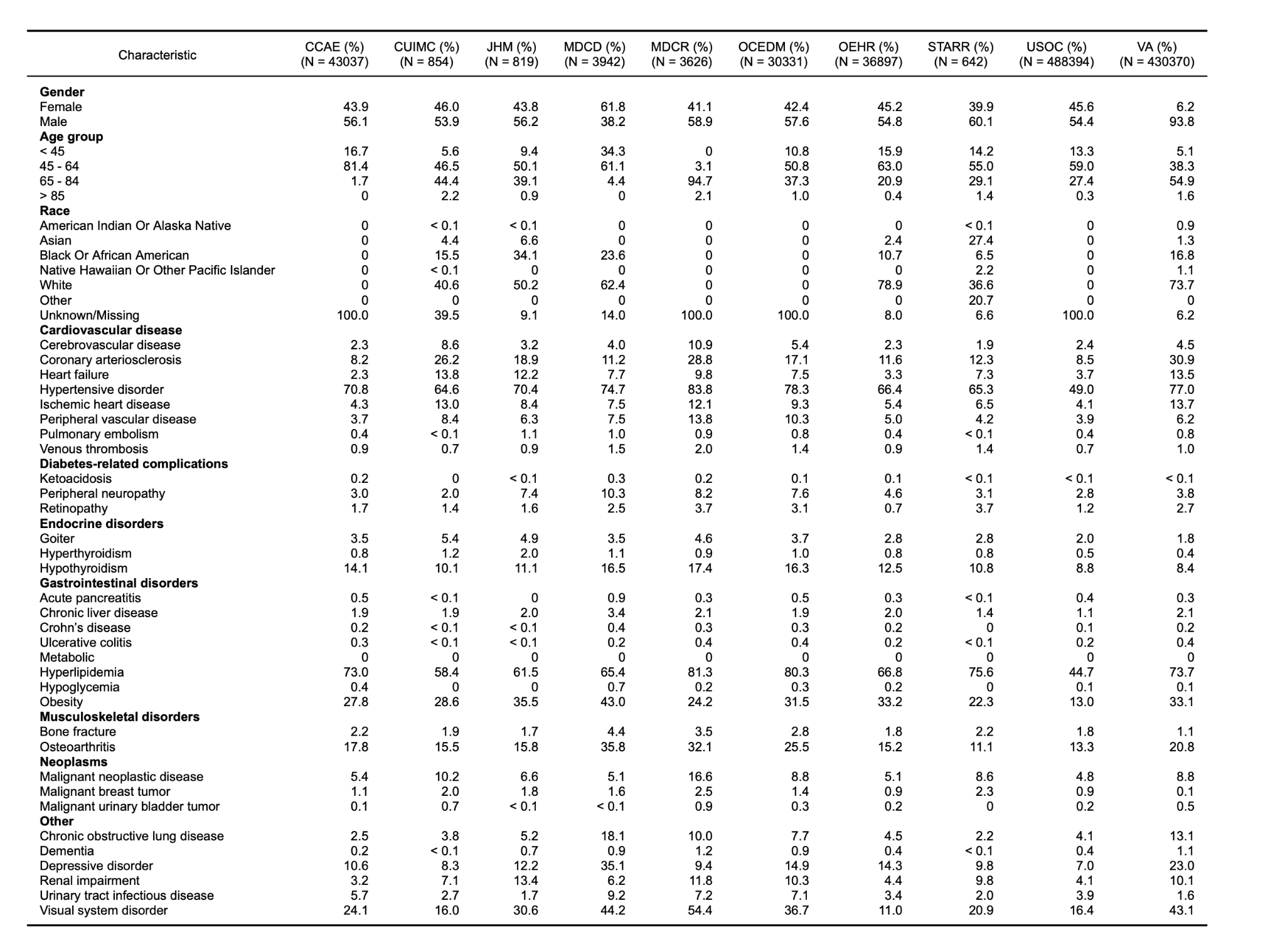

**Abbreviations:** CCAE: IBM MarketScan® Commercial Claims and Encounters Data (CCAE), CUIMC: Columbia University Irving Medical Center, JHM: Johns Hopkins Medicine, MDCD: IBM Health MarketScan® Multi-State Medicaid Database, MDCR: IBM Health MarketScan® Medicare Supplemental and Coordination of Benefits Database, OCEDM: Optum Clinformatics Extended Data Mart - Date of Death (DOD), OEHR: Optum© de-identified Electronic Health Record Dataset, STARR: Stanford Medicine, USOC: US Open Claims

**Supplemental Table S6 |** **Baseline Characteristics and Clinical Covariates in Patients Using Dipeptidyl Peptidase-4 Inhibitors as Second-Line Antihyperglycemic Agents in US Databases**

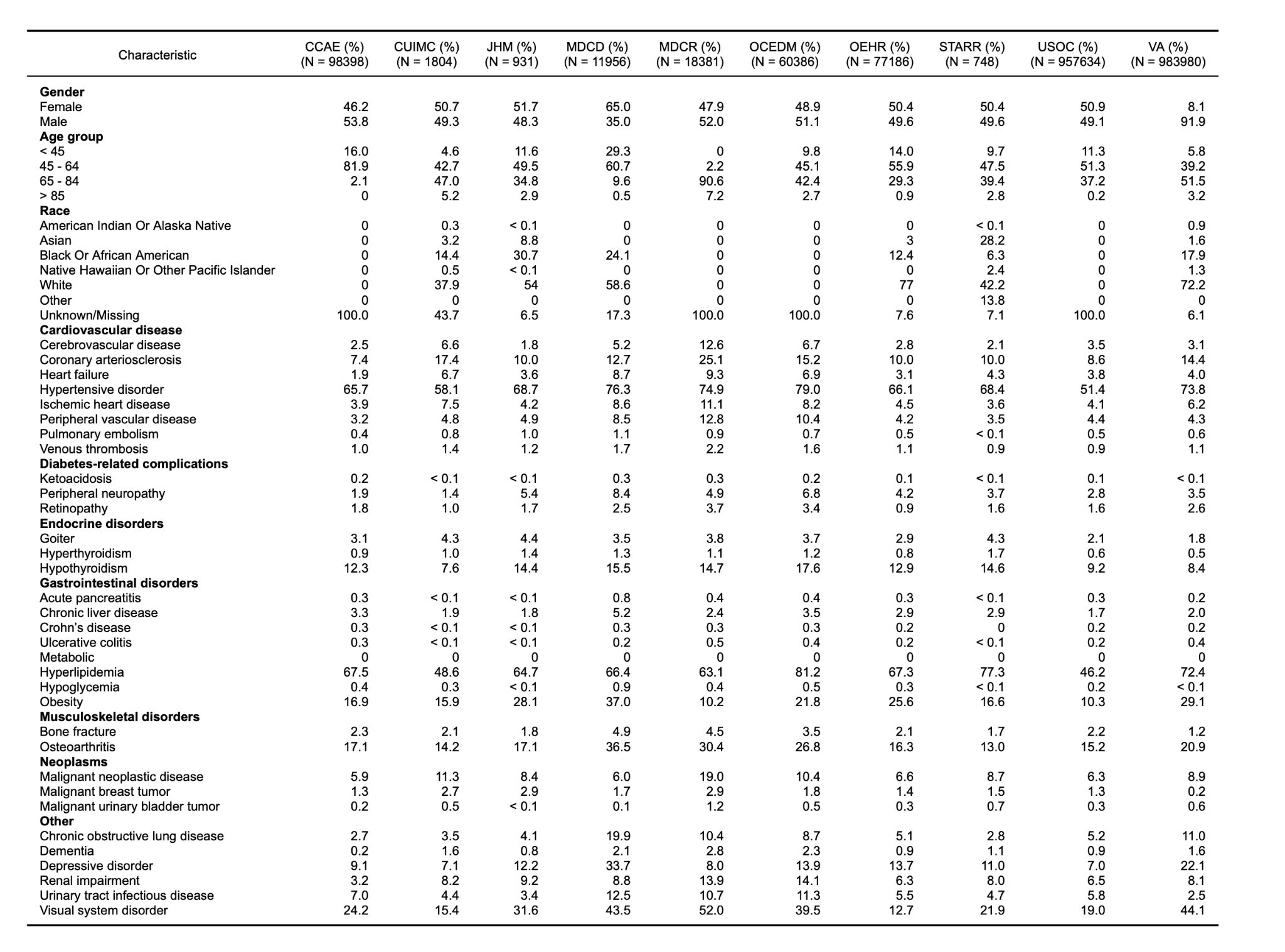

**Abbreviations:** CCAE: IBM MarketScan® Commercial Claims and Encounters Data (CCAE), CUIMC: Columbia University Irving Medical Center, JHM: Johns Hopkins Medicine, MDCD: IBM Health MarketScan® Multi-State Medicaid Database, MDCR: IBM Health MarketScan® Medicare Supplemental and Coordination of Benefits Database, OCEDM: Optum Clinformatics Extended Data Mart - Date of Death (DOD), OEHR: Optum© de-identified Electronic Health Record Dataset, STARR: Stanford Medicine, USOC: US Open Claims

**Supplemental Table S7 |** **Baseline Characteristics and Clinical Covariates in Patients Using Sulfonylureas as Second-Line Antihyperglycemic Agents in US Databases**

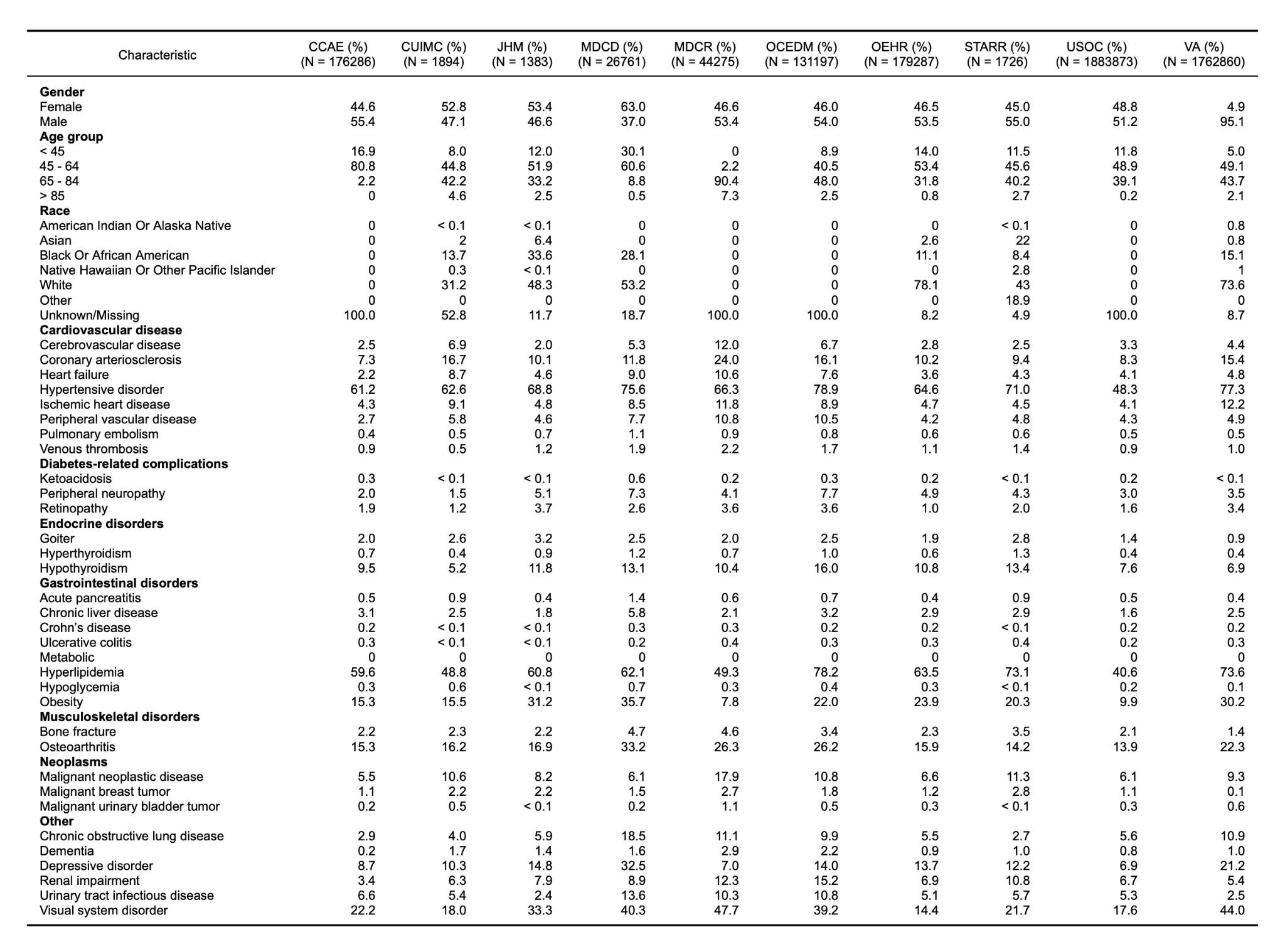

**Abbreviations:** CCAE: IBM MarketScan® Commercial Claims and Encounters Data (CCAE), CUIMC: Columbia University Irving Medical Center, JHM: Johns Hopkins Medicine, MDCD: IBM Health MarketScan® Multi-State Medicaid Database, MDCR: IBM Health MarketScan® Medicare Supplemental and Coordination of Benefits Database, OCEDM: Optum Clinformatics Extended Data Mart - Date of Death (DOD), OEHR: Optum© de-identified Electronic Health Record Dataset, STARR: Stanford Medicine, USOC: US Open Claims
**Supplemental Table S8 | Baseline Characteristics and Clinical Covariates in Patients Using Glucagon-like Peptide-1 Receptor Agonists as Second-Line Antihyperglycemic Agents in non-United States Databases**

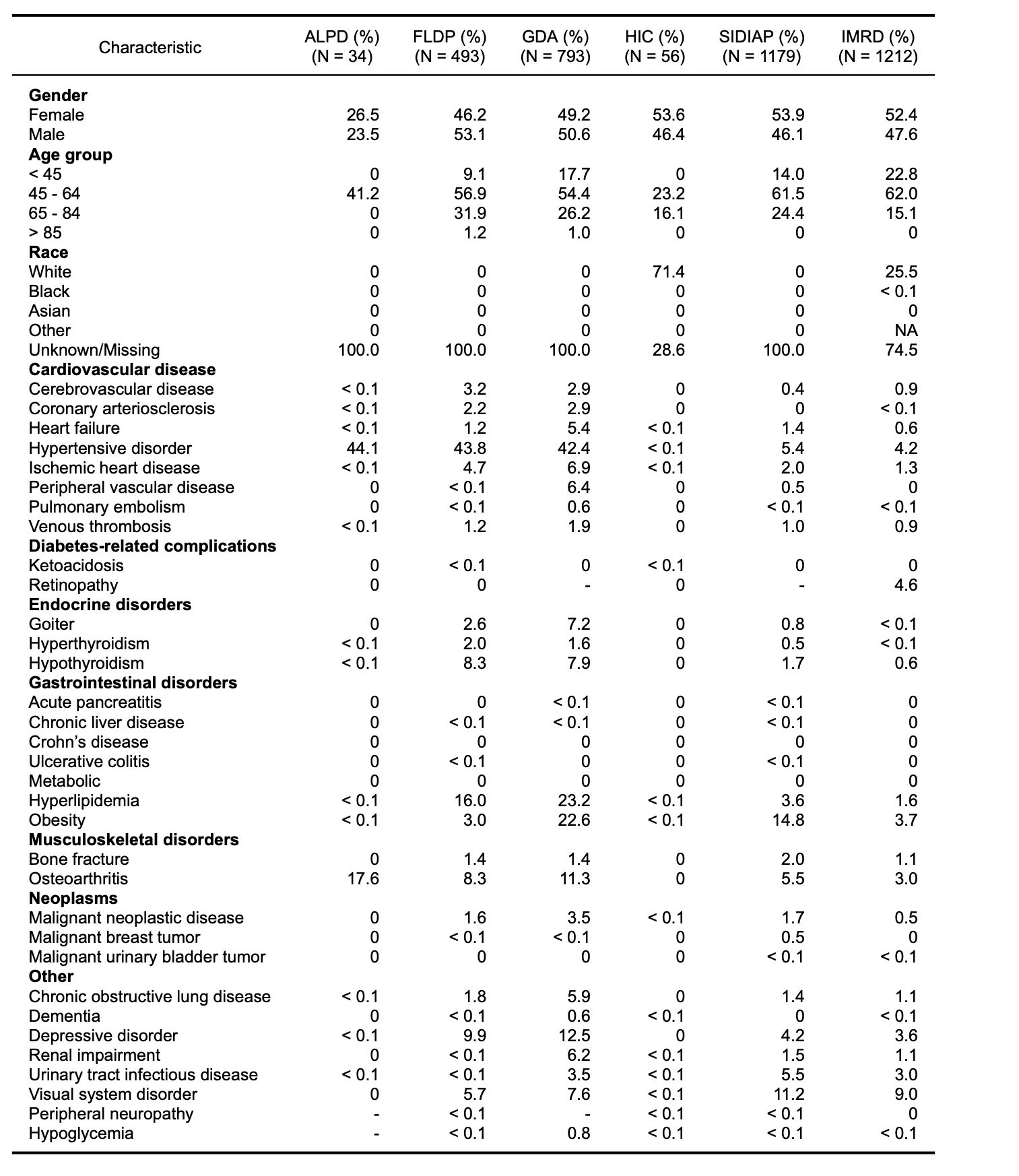

**Abbreviations:** ALPD: Australia Longitudinal Patient Database Practice Profile, FLPD: France Longitudinal Patient Database, GDA: Germany Disease Analyser, HIC: Health Informatics Centre at the University of Dundee, IMRD: United Kingdom-IQVIA Medical Research Data SIDIAP: Information System for Research in Primary Care

**Supplemental Table S9 | Baseline Characteristics and Clinical Covariates in Patients Using Sodium-Glucose Cotransporter 2 Inhibitors as Second-Line Antihyperglycemic Agents in non-United States Databases**

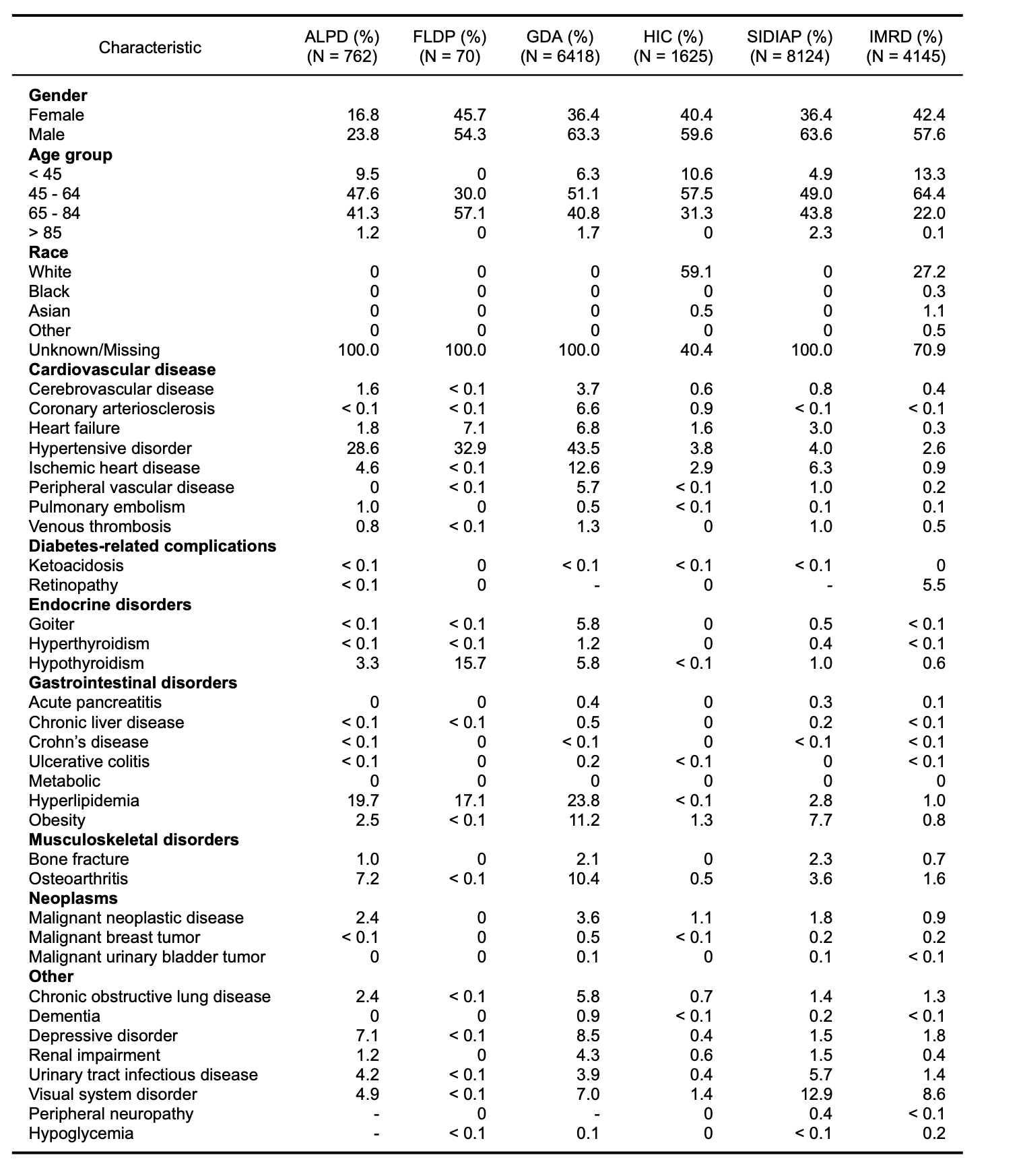

**Abbreviations:** ALPD: Australia Longitudinal Patient Database Practice Profile, FLPD: France Longitudinal Patient Database, GDA: Germany Disease Analyser, HIC: Health Informatics Centre at the University of Dundee, IMRD: United Kingdom-IQVIA Medical Research Data SIDIAP: Information System for Research in Primary Care

**Supplemental Table S10 |** **Baseline Characteristics and Clinical Covariates in Patients Using Dipeptidyl Peptidase-4 Inhibitors as Second-Line Antihyperglycemic Agents in non-United States Databases**

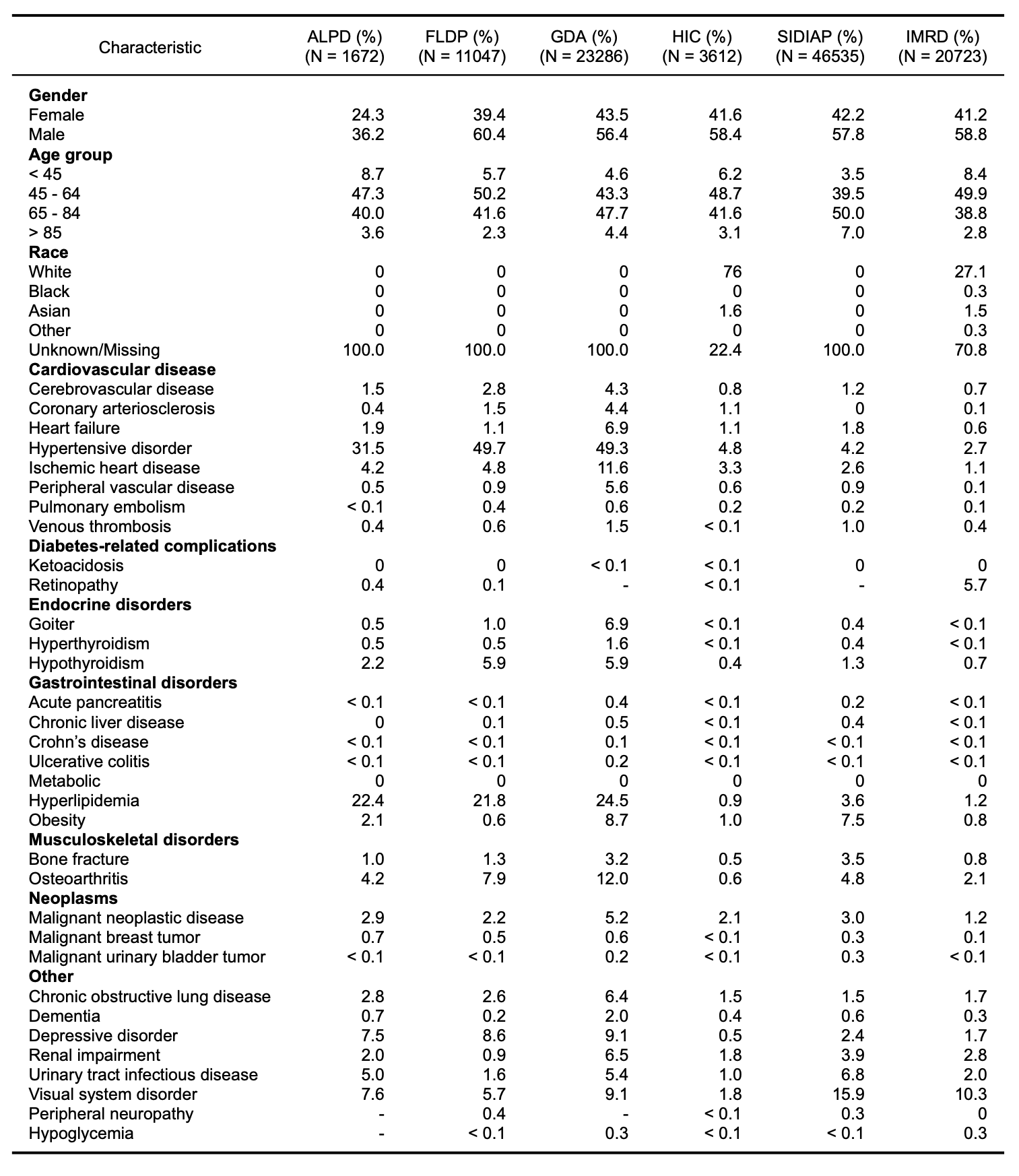

**Abbreviations:** ALPD: Australia Longitudinal Patient Database Practice Profile, FLPD: France Longitudinal Patient Database, GDA: Germany Disease Analyser, HIC: Health Informatics Centre at the University of Dundee, IMRD: United Kingdom-IQVIA Medical Research Data SIDIAP: Information System for Research in Primary Care

**Supplemental Table S11 |** **Baseline Characteristics and Clinical Covariates in Patients Using Sulfonylureas as Second-Line Antihyperglycemic Agents in non-United States Databases**

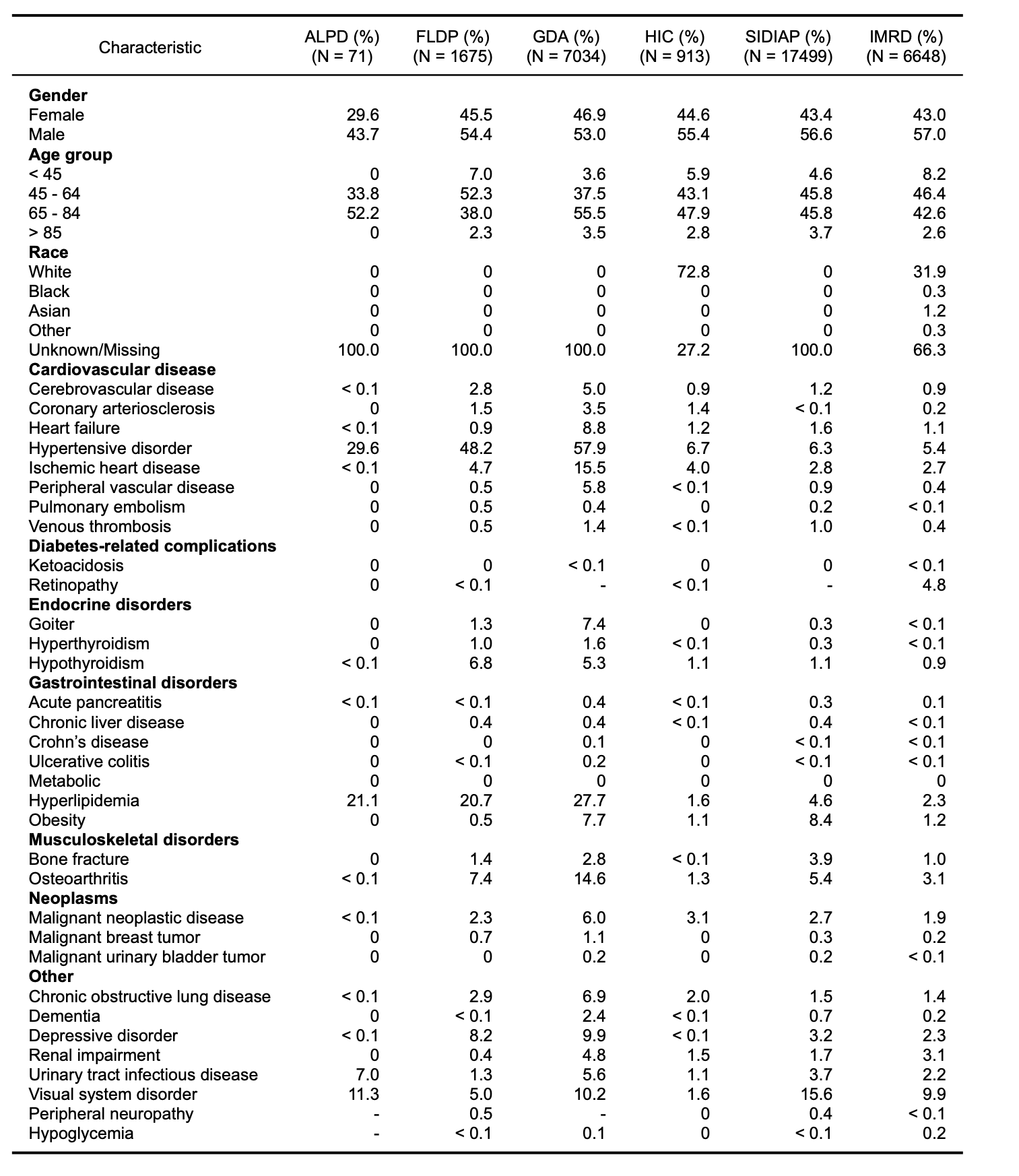

**Abbreviations:** ALPD: Australia Longitudinal Patient Database Practice Profile, FLPD: France Longitudinal Patient Database, GDA: Germany Disease Analyser, HIC: Health Informatics Centre at the University of Dundee, IMRD: United Kingdom-IQVIA Medical Research Data SIDIAP: Information System for Research in Primary Care

**Supplemental Table S12 |** **Annualized Change in the Incident Use of Dipeptidyl Peptidase-4 Inhibitors** **for Patients with Established Cardiovascular Disease and Patients without Established Cardiovascular Disease**

| **Data Source** | **Slope for Patients with CVD** | **Slope for Patients without CVD** | **P-value for Slope Difference** |
| --- | --- | --- | --- |
| **US National Databases** | | | |
| CCAE | -0.75% (-1.17 to -0.33) | -1.9% (-2.99 to -0.82) | 0.025 |
| MDCD | -0.07% (-0.36 to 0.22) | -0.93% (-1.56 to -0.31) | 0.007 |
| MDCR | -3.92% (-11.75 to 3.92) | -3.92% (-8.49 to 0.65) | 0.999 |
| OCEDM | 0.86% (0.43 to 1.3) | -0.69% (-1.56 to 0.19) | 0.002 |
| OEHR | 1.16% (0.65 to 1.67) | 2.21% (1.39 to 3.03) | 0.016 |
| USOC | 0.07% (-0.45 to 0.6) | -0.32% (-1.35 to 0.71) | 0.373 |
| **US Health System Databases** | | | |
| CUIMC | -0.17% (-1.3 to 0.96) | -0.18% (-0.42 to 0.06) | 0.993 |
| JHM | 0.23% (-0.37 to 0.82) | -0.31% (-3.97 to 3.35) | 0.661 |
| STARR | 0.04% (-0.35 to 0.42) | 0.12% (-0.41 to 0.66) | 0.716 |
| VA | 6.93% (5.54 to 8.32) | 12.65% (10.09 to 15.21) | 0.001 |
| **Non-US Databases** | | | |
| ALPD | -0.4% (-2.05 to 1.25) | 9.02% (1.67 to 16.37) | 0.007 |
| FLPD | 1.18% (0.53 to 1.83) | 6.7% (3.08 to 10.33) | 0.003 |
| GDA | 3.38% (1.62 to 5.14) | 4.34% (2.1 to 6.58) | 0.376 |
| HIC | -0.35% (-0.99 to 0.28) | -0.19% (-1.64 to 1.25) | 0.782 |
| HKHA | 7.26% (-23.68 to 38.2) | 12% (-55.53 to 79.52) | 0.503 |
| IMRD | -0.12% (-0.46 to 0.23) | 0.67% (-2.82 to 4.16) | 0.39 |
| SIDIAP | 1.28% (-0.56 to 3.12) | 6.72% (-2.29 to 15.73) | 0.139 |

**Abbreviations:** ALPD - Australia Longitudinal Patient Database Practice Profile, CCAE - IBM MarketScan® Commercial Claims and Encounters Data (CCAE), CUIMC - Columbia University Irving Medical Center, FLPD - France Longitudinal Patient Database, GDA - Germany Disease Analyser, HIC - Health Informatics Centre at the University of Dundee, HKHA - Hong Kong Hospital Authority, IMRD - UK-IQVIA Medical Research Data, JHM - Johns Hopkins Medicine, MDCD - IBM Health MarketScan® Multi-State Medicaid Database, MDCR - IBM Health MarketScan® Medicare Supplemental and Coordination of Benefits Database, OCEDM - Optum Clinformatics Extended Data Mart - Date of Death (DOD), OEHR - Optum© de-identified Electronic Health Record Dataset, SIDIAP - Information System for Research in Primary Care, STARR - Stanford Medicine, USOC - United States Open Claims, VA - Department of Veterans Affairs Healthcare System

**Supplemental Table S13 |** **Annualized Change in the Incident Use of Sulfonylureas for Patients with Established Cardiovascular Disease and Patients without Established Cardiovascular Disease**

| **Data Source** | **Slope for Patients with CVD** | **Slope for Patients without CVD** | **P-value for Slope Difference** |
| --- | --- | --- | --- |
| **US National Databases** | | | |
| CCAE | -0.29% (-0.97 to 0.39) | -1.58% (-3.21 to 0.05) | 0.078 |
| MDCD | -0.14% (-0.57 to 0.29) | -1.67% (-2.38 to -0.97) | 0.001 |
| MDCR | -5.75% (-15.17 to 3.66) | -3.94% (-11.27 to 3.39) | 0.685 |
| OCEDM | 2.47% (1.54 to 3.4) | 0.53% (-1.07 to 2.12) | 0.019 |
| OEHR | 2.17% (1.36 to 2.99) | 3.97% (1.46 to 6.49) | 0.096 |
| USOC | 1.13% (-0.1 to 2.36) | 1.63% (-1.15 to 4.41) | 0.663 |
| **US Health System Databases** | | | |
| CUIMC | -0.46% (-0.86 to -0.06) | -0.81% (-1.3 to -0.33) | 0.159 |
| JHM | -0.13% (-1.5 to 1.25) | -0.99% (-5.02 to 3.05) | 0.544 |
| STARR | -0.26% (-0.69 to 0.17) | -1.07% (-1.92 to -0.22) | 0.046 |
| VA | -3.07% (-8.1 to 1.96) | -3.09% (-9.49 to 3.3) | 0.994 |
| **Non-US Databases** | | | |
| ALPD | 0% (0 to 0) | -0.05% (-1.24 to 1.13) | 0.892 |
| FLPD | 0.07% (-0.07 to 0.2) | 0.12% (-0.46 to 0.7) | 0.797 |
| GDA | -0.16% (-0.28 to -0.03) | -0.08% (-0.29 to 0.14) | 0.389 |
| HIC | -0.19% (-0.36 to -0.01) | -0.62% (-1 to -0.25) | 0.019 |
| HKHA | 2.89% (-30.37 to 36.14) | 0.63% (-58.28 to 59.53) | 0.712 |
| IMRD | -0.41% (-0.83 to 0) | -0.03% (-0.79 to 0.73) | 0.13 |
| SIDIAP | -0.21% (-0.48 to 0.06) | -1.32% (-2.4 to -0.25) | 0.023 |

**Abbreviations:** ALPD - Australia Longitudinal Patient Database Practice Profile, CCAE - IBM MarketScan® Commercial Claims and Encounters Data (CCAE), CUIMC - Columbia University Irving Medical Center, FLPD - France Longitudinal Patient Database, GDA - Germany Disease Analyser, HIC - Health Informatics Centre at the University of Dundee, HKHA - Hong Kong Hospital Authority, IMRD - UK-IQVIA Medical Research Data, JHM - Johns Hopkins Medicine, MDCD - IBM Health MarketScan® Multi-State Medicaid Database, MDCR - IBM Health MarketScan® Medicare Supplemental and Coordination of Benefits Database, OCEDM - Optum Clinformatics Extended Data Mart - Date of Death (DOD), OEHR - Optum© de-identified Electronic Health Record Dataset, SIDIAP - Information System for Research in Primary Care, STARR - Stanford Medicine, USOC - United States Open Claims, VA - Department of Veterans Affairs Healthcare System

**Supplemental Figure S1 |** **Proportional Incident Use of Second-Line Antihyperglycemic Agents in United States National Databases in 2020**
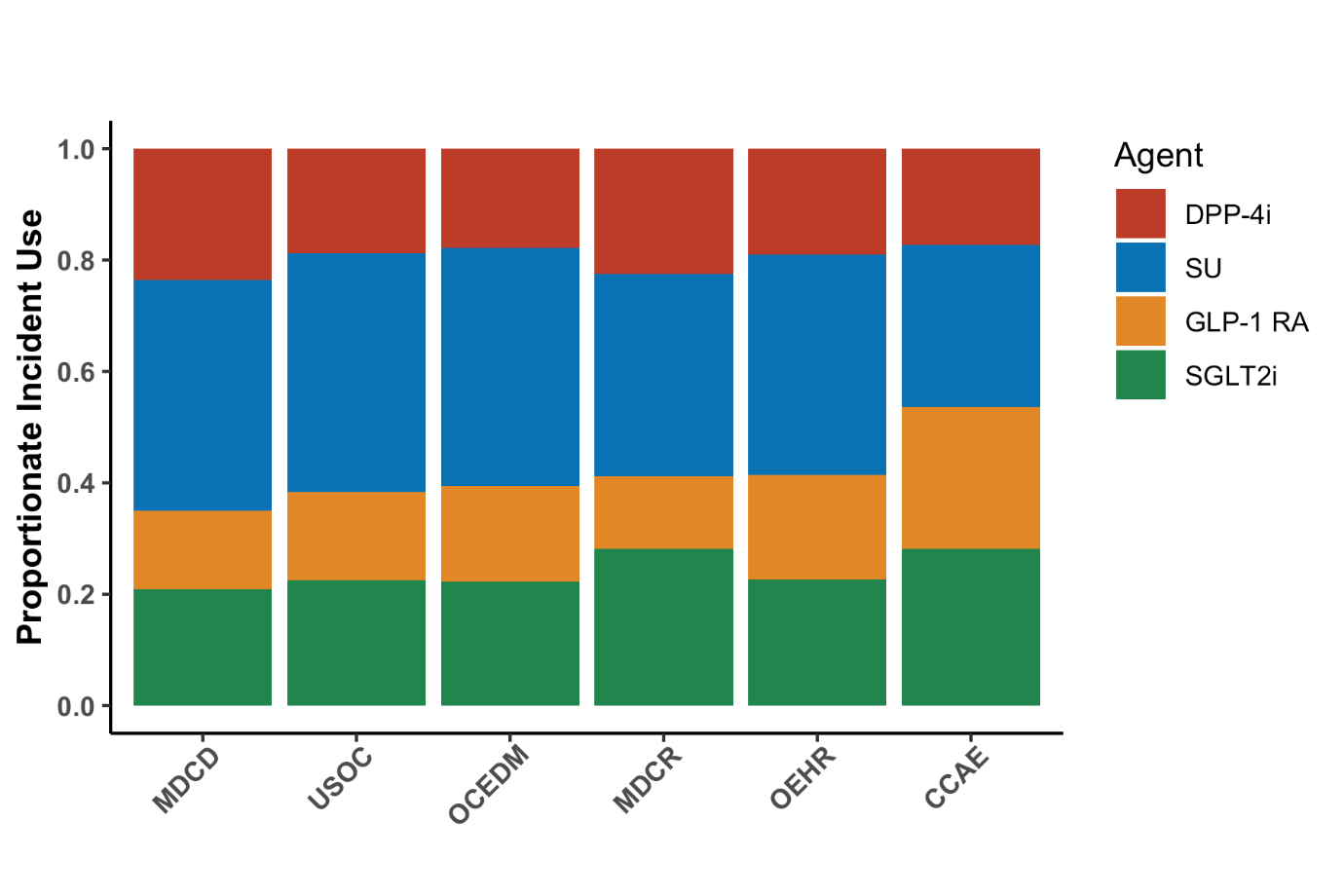

**Abbreviations:** CCAE - IBM MarketScan® Commercial Claims and Encounters Data (CCAE), DPP-4i - Dipeptidyl Peptidase-4 Inhibitors, GLP-1 RA - Glucagon-like Peptide-1 Receptor Agonist, MDCD - IBM Health MarketScan® Multi-State Medicaid Database, MDCR - IBM Health MarketScan® Medicare Supplemental and Coordination of Benefits Database, OCEDM - Optum Clinformatics Extended Data Mart - Date of Death (DOD), OEHR - Optum© de-identified Electronic Health Record Dataset, SGLT2i - Sodium-Glucose Cotransporter 2 Inhibitor, SU - Sulfonylurea, USOC - United States Open Claims

**Supplemental Figure S2 |** **Proportional Incident Use of Second-Line Antihyperglycemic Agents in United States Health System Databases in 2020**

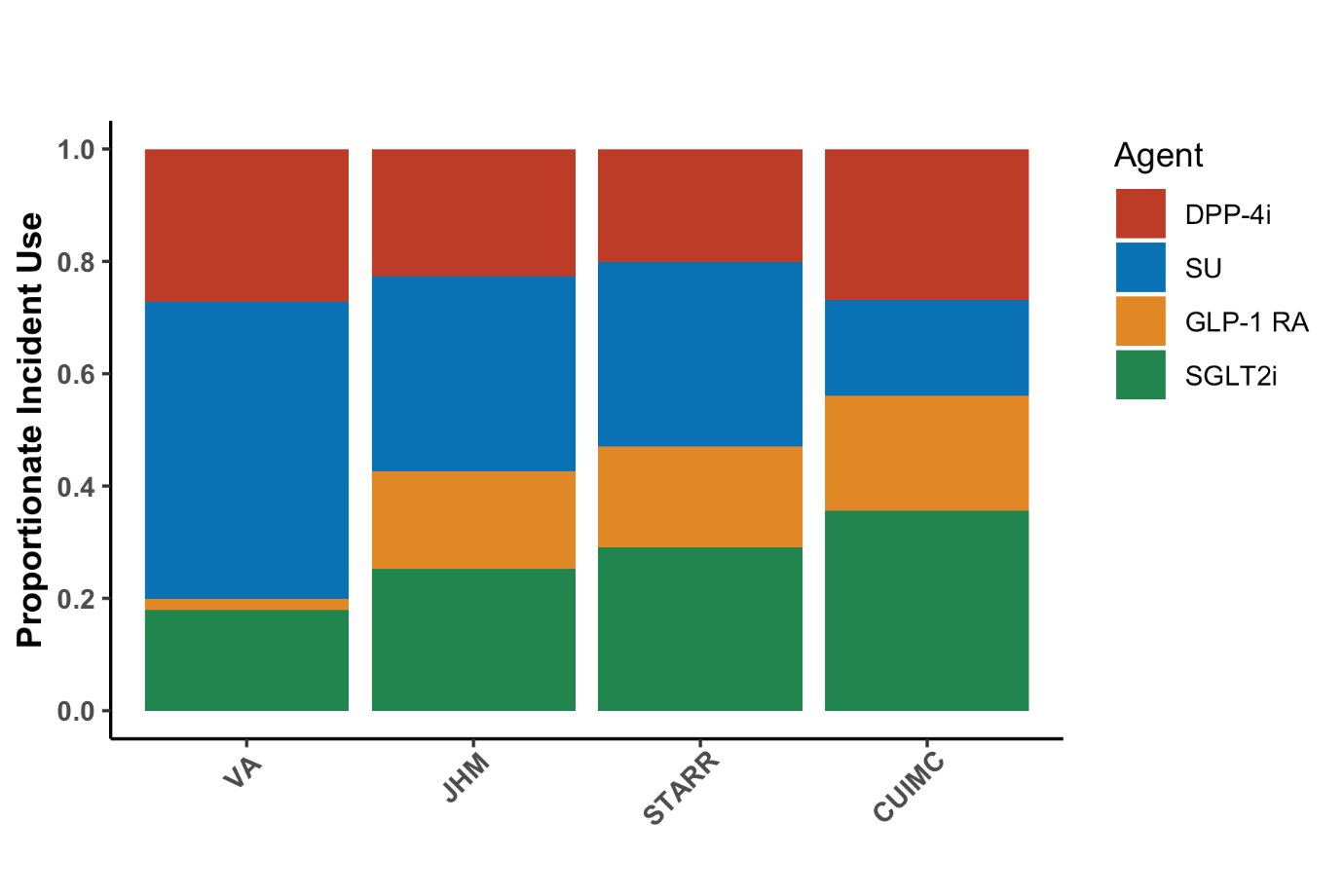

**Abbreviations:** CUIMC - Columbia University Irving Medical Center, DPP-4i - Dipeptidyl Peptidase-4 Inhibitors, GLP-1 RA - Glucagon-like Peptide-1 Receptor Agonist, JHM - Johns Hopkins Medicine, SGLT2i - Sodium-Glucose Cotransporter 2 Inhibitor, STARR - Stanford Medicine, SU - Sulfonylurea, VA - Department of Veterans Affairs Healthcare System

**Supplemental Figure S3 |** **Proportional Incident Use of Second-Line Antihyperglycemic Agents in non-United States Databases in 2020**

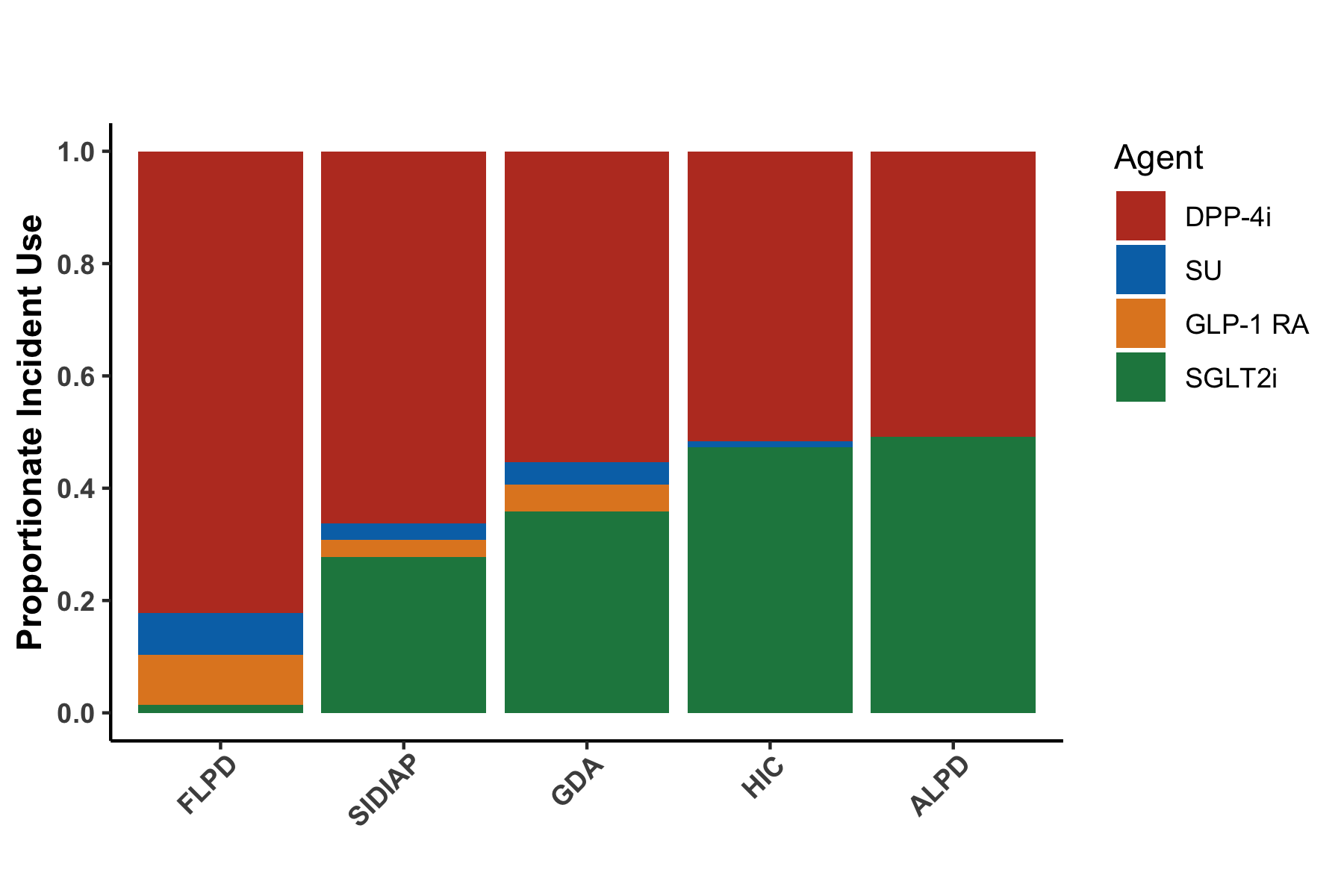

**Abbreviations:** ALPD - Australia Longitudinal Patient Database Practice Profile, DPP-4i - Dipeptidyl Peptidase-4 Inhibitors, FLPD - France Longitudinal Patient Database, GDA - Germany Disease Analyser, GLP-1 RA - Glucagon-like Peptide-1 Receptor Agonist, HIC - Health Informatics Centre at the University of Dundee, HKHA - Hong Kong Hospital Authority, SGLT2i - Sodium-Glucose Cotransporter 2 Inhibitor, SIDIAP - Information System for Research in Primary Care, SU - Sulfonylurea

**Supplemental Figure S4 | Yearly Trends in Proportional Incident Use of Second-Line Antihyperglycemic Agents in US National Databases**

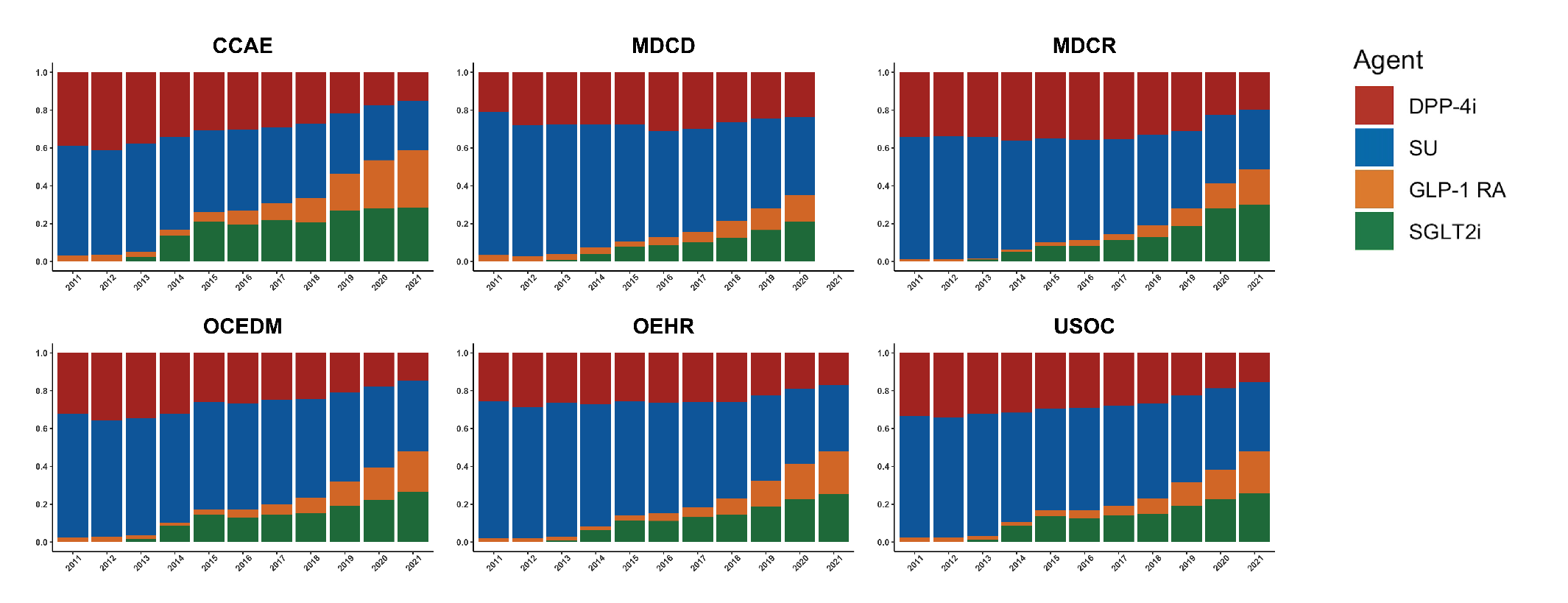

Y-axes are the proportion of each drug used among those initiating a second-line T2DM drug in a calendar year, and X-axes represent calendar years.

**Abbreviations:** Abbreviations: CCAE - IBM MarketScan® Commercial Claims and Encounters Data (CCAE), DPP-4i - Dipeptidyl Peptidase-4 Inhibitors, GLP-1 RA - Glucagon-like Peptide-1 Receptor Agonist, MDCD - IBM Health MarketScan® Multi-State Medicaid Database, MDCR - IBM Health MarketScan® Medicare Supplemental and Coordination of Benefits Database, OCEDM - Optum Clinformatics Extended Data Mart - Date of Death (DOD), OEHR - Optum© de-identified Electronic Health Record Dataset, SGLT2i - Sodium-Glucose Cotransporter 2 Inhibitor, SU - Sulfonylurea, USOC - United States Open Claims

**Supplemental Figure S5 | Yearly Trends in Proportional Incident Use of Second-Line Antihyperglycemic Agents in US Health System Databases**

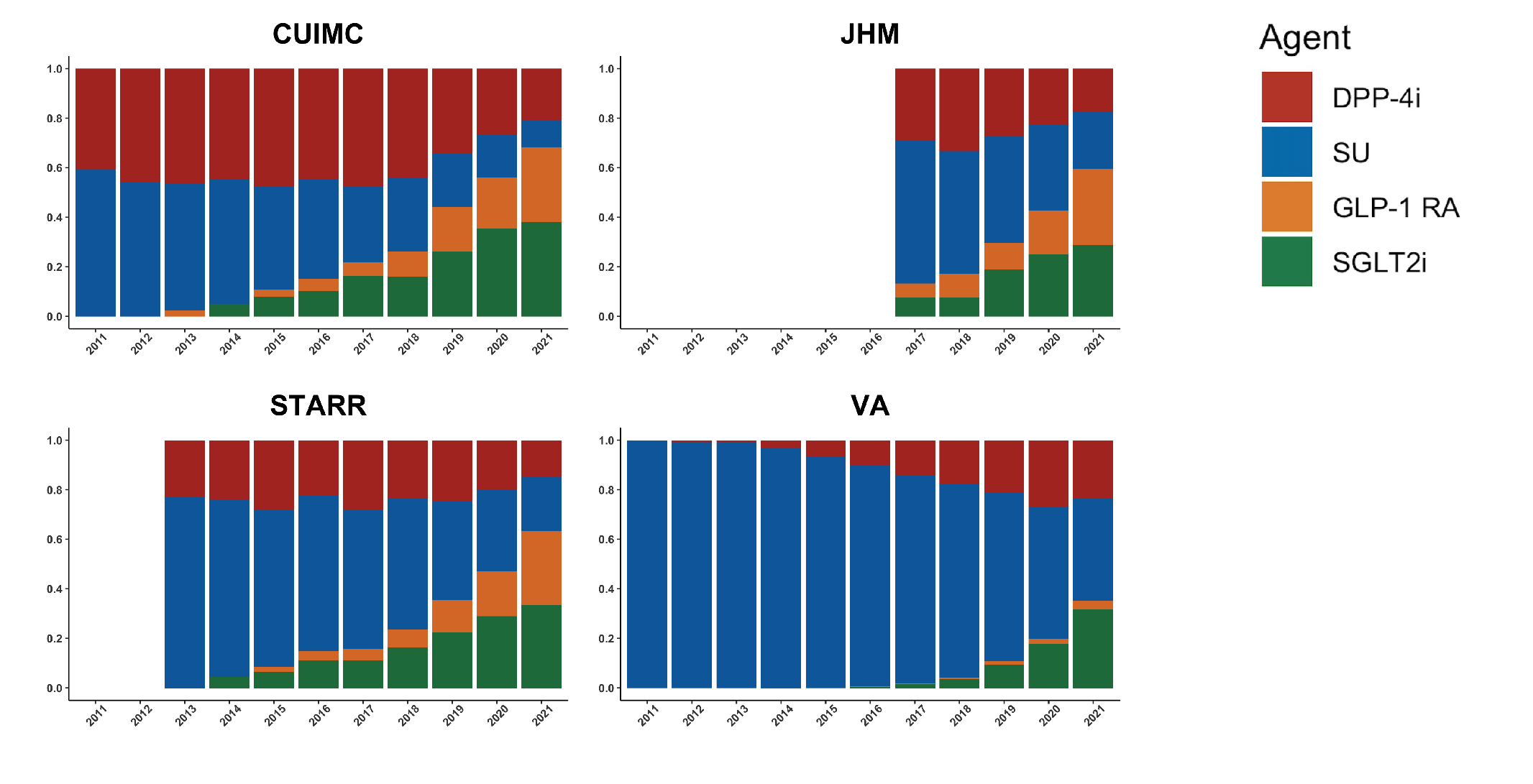

Y-axes are the proportion of each drug used among those initiating a second-line T2DM drug in a calendar year, and X-axes represent calendar years.

**Abbreviations:** Abbreviations: CUIMC - Columbia University Irving Medical Center, DPP-4i - Dipeptidyl Peptidase-4 Inhibitors, GLP-1 RA - Glucagon-like Peptide-1 Receptor Agonist, JHM - Johns Hopkins Medicine, SGLT2i - Sodium-Glucose Cotransporter 2 Inhibitor, STARR - Stanford Medicine, SU - Sulfonylurea, VA - Department of Veterans Affairs Healthcare System

**Supplemental Figure S6 |** **Yearly Trends in Proportional Incident Use of Second Line Antihyperglycemic Agents in non-United States Databases**

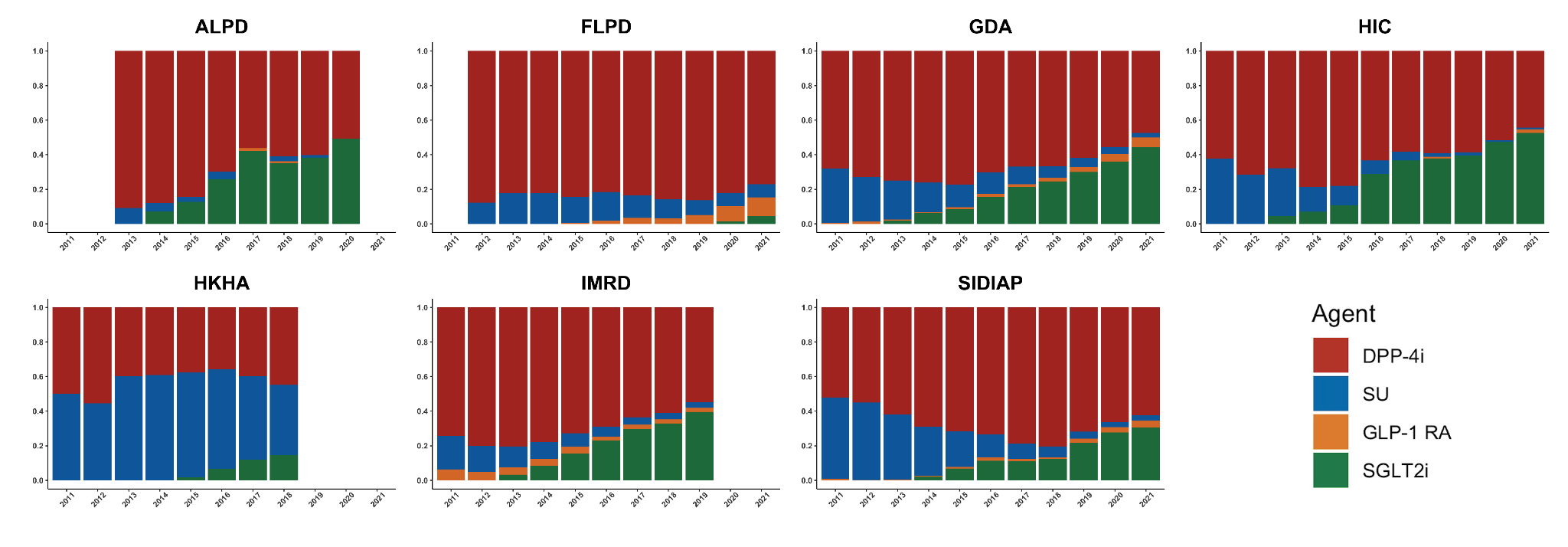

Y-axes are the proportion of each drug used among those initiating a second-line T2DM drug in a calendar year, and X-axes represent calendar years. **Abbreviations:** ALPD - Australia Longitudinal Patient Database Practice Profile, DPP-4i - Dipeptidyl Peptidase-4 Inhibitors, FLPD - France Longitudinal Patient Database, GDA - Germany Disease Analyser, GLP-1 RA - Glucagon-like Peptide-1 Receptor Agonist, HIC - Health Informatics Centre at the University of Dundee, HKHA - Hong Kong Hospital Authority, IMRD - UK-IQVIA Medical Research Data, SGLT2i - Sodium-Glucose Cotransporter 2 Inhibitor, SIDIAP - Information System for Research in Primary Care, SU - Sulfonylurea
